# SARS-CoV-2 Damages Cardiomyocytes Mitochondria and Implicates Long COVID-associated Cardiovascular Manifestations

**DOI:** 10.1101/2024.08.18.24311961

**Authors:** Wenliang Che, Shuai Guo, Yanqun Wang, Xiaohua Wan, Bingyu Tan, Hailing Li, Jiasuer Alifu, Mengyun Zhu, Cesong Chen, Peiyao Li, Zhaoyong Zhang, Yiliang Wang, Xiaohan Huang, Xinsheng Wang, Jian Zhu, Xijiang Pan, Fa Zhang, Peiyi Wang, Jincun Zhao, Yawei Xu, Zheng Liu

**Author notes:** These authors contributed equally (Wenliang Che, Shuai Guo, Yanqun Wang). Correspondence (Z.L.), (Y.X.), (J.Z.).

## Abstract

Our study investigates the persistent cardiovascular symptoms observed in individuals long after contracting SARS-CoV-2, a condition commonly referred to as “Long COVID”, which has significantly affected millions globally. We meticulously describe the cardiovascular outcomes in five patients, encompassing a range of severe conditions such as sudden cardiac death during exercise, coronary atherosclerotic heart disease, acute inferior myocardial infarction, and acute myocarditis. All five patients were diagnosed with myocarditis, confirmed through endomyocardial biopsy and histochemical staining, which identified inflammatory cell infiltration in their heart tissue. Crucially, electron microscopy revealed widespread mitochondrial vacuolations and the presence of myofilament degradation within the cardiomyocytes of these patients. These findings were mirrored in SARS-CoV-2-infected mice, suggesting a potential underlying cellular mechanism for the cardiac effects associated with Long COVID. Our report sheds light on the cardiovascular implications of Long COVID and underscores the importance of further research to understand its cellular underpinnings.

## INTRODUCTION

Severe acute respiratory syndrome coronavirus 2 (SARS-COV-2), the causative agent of the coronavirus disease 2019 (COVID-19) pandemic, emerged in late 2019 and rapidly spread around the world. By the end of June 2023, there have been more than 775 million confirmed cases of COVID-19, with over 7.0 million fatalities worldwide (https://covid19.who.int/). While primarily affecting the respiratory system, SARS-CoV-2 is also linked to multi-organ dysfunctions, with a notable impact on the cardiovascular system. The virus utilizes the angiotensin-converting enzyme 2 (ACE2) receptor, which is abundantly expressed in cardiomyocytes, to enter human cells, thereby underlining a pathway to cardiac involvement (Shang et al., 2020; Yang et al., 2021). Clinical reports have underscored the cardiovascular complications associated with COVID-19, including myocardial injury, arrhythmias, and severe clinical outcomes (Guo et al., 2020; Huang et al., 2020; Wang et al., 2020).

Beyond the acute phase, a considerable number of recovered patients experience long-term cardiovascular issues, such as cerebrovascular disorders, dysrhythmias, and myocarditis, contributing to the spectrum of “Post-acute COVID-19 Syndrome” or “Long COVID” (Nalbandian et al., 2021; Xie et al., 2022). Long COVID is broadly defined as signs, symptoms, and conditions that continue or develop after initial COVID-19 or SARS-CoV-2 infection. The signs, symptoms, and conditions persist for four weeks or more after the initial phase of infection. This working definition was developed by the U.S. Department of Health and Human Services in collaboration with CDC, NIH, and medical societies (https://www.covid.gov/longcovid/). A recent longitudinal cohort study has reported that approximately one in eight patients, or 12.7%, have symptoms attributed to COVID-19, and that chest pain is one of the most commonly reported symptoms of Long COVID (Ballering et al., 2022).

The specific pathophysiology underlying cardiovascular manifestations in Long COVID remains largely unexplored. Our investigation provides histopathological and electron microscopic analyses from endomyocardial biopsies of five patients who experienced various cardiovascular outcomes, such as acute myocarditis and myocardial fibrosis, within one to three months post-COVID-19 recovery. Findings include inflammatory cell infiltration, myofilament degradation, and notably, extensive mitochondrial vacuolations and lipofuscin accumulation within cardiomyocytes. These observations suggest a profound impact of SARS-CoV-2 on mitochondrial integrity, potentially elucidating a cellular mechanism behind the cardiovascular sequelae of Long COVID.

## RESULTS

### Ultrastructural disorder in cardiomyocytes in a patient suffer sudden cardiac death

A 30’s male suffered a syncope during exercise, leading to lost consciousness, cessation of breathing, and absence of carotid pulse. Immediate intervention with two cycles of cardiopulmonary resuscitation (CPR) and automated external defibrillator (AED) deployment successfully restored his heartbeat and breathing, although he remained unconscious. Upon regaining consciousness en route to emergency room, the patient had limited recollection of the event. Initial assessment in the coronary care unit (CCU) for further diagnosis. The electrocardiogram (ECG) at the time of the AED discharge indicated ventricular fibrillation, prompted further investigation (**Figure S1** and **Fig. 1A**).

**Figure 1.**
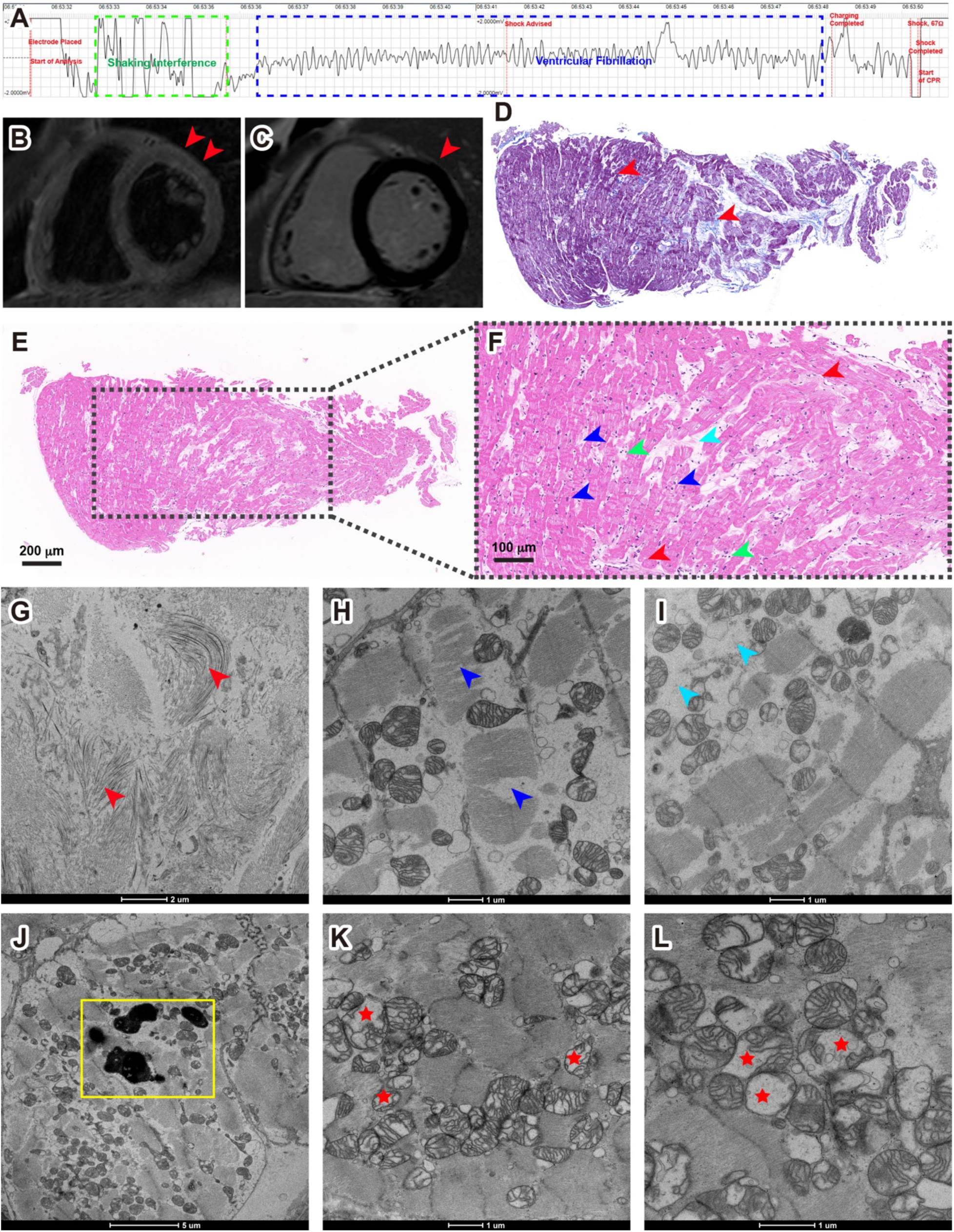
Spectrum of myocarditis in COVID-19 Patient #1. (A) ECG recordings retrieved from the AED, shows coarse ventricular fibrillation before the shock (blue frame). (B) CMR imaging of the patient, both T2-weighted sequences (T2WI) and short inversion time inversion recovery sequences (STIR) showed no myocardial edema in the mid short-axis slices. (C) a late enhancement was not observed in late gadolinium enhancement (LGE) sequences of the mid short-axis slices in the myocardium. Red arrowheads indicated suspected myocardial edema under the epicardium of the anterior wall (basal part), without significant delayed enhancement. (D) Masson’s trichrome staining displayed interstitial collagen fiber deposits (red arrowheads). (E) HE analysis shows an abnormal structure in the myocardium. (F) zoomed view, shows interstitial collagen fiber accumulation (red arrowheads), loss of integrity of myofibrillar bundles (blue arrowheads), and edema and necrosis of myofibrillar bundles (cyan arrowhead); Myofibrillar interstitial infiltration with inflammatory cells is also observed (green arrowheads). (G) electron micrographs of the biopsy sample from the patient, showing local myofibrillar fibrosis and proliferation of fibroblasts (red arrowheads); (H) loss of integrity of myofibrillar bundles (blue arrowheads); (I) necrosis of myofibrillar bundles (cyan arrowheads); (J) lipofuscin granules in the cardiomyocytes (yellow frames); (K) and (L) a significant amount of vacuolations in the mitochondria (red stars).

The patient has recent recovered from COVID-19, 37 days prior to the incident, was notable in the absence of any chronic health conditions or familial cardiovascular disease history. Comprehensive laboratory assessments revealed generally normal results, with specific abnormalities including elevated cardiac biomarkers (Troponin T, creatine kinase-MB, and myoglobin) and abnormal neutrophil counts indicative of myocardial injury and inflammation, respectively (**Table S1**). All pathogen tests, including SARS-CoV-2 and Coxsackie virus, were negative. Additional investigations resulted in no signs of rheumatic heart disease, autoimmune disease, and tumors that may cause myocardial injury. We considered possible viral myocarditis (VMC), however, only suspicious myocardial edema in the anterior subepicardial with no significant delayed enhancement in cardiac magnetic resonance (CMR, **Figure 1B** and **1C**). Coronary angiography (CAG) confirmed unobstructed coronary arteries. Further diagnostic tools, including ECG, echocardiography (Echo), 24-h dynamic electrocardiogram, endocardial electrophysiological examination, and genetic analysis, revealed no abnormalities.

Endomyocardial biopsy remains the gold standard mode of investigation for diagnosing many cardiac conditions, including suspected myocarditis, heart failure of unknown etiology, cardiomyopathy, arrhythmia, heart transplant rejection and secondary involvement by systemic diseases (Cunningham et al., 2006). To further clarify the pathogeny of the patient, an endomyocardial biopsy was performed. The biopsy sample underwent immunohistochemical analysis to characterize inflammatory cell infiltration by using the following antibodies: CD3, CD4, CD8, CD9, CD20, and CD68. Only CD68 showed weekly positive, which indicated a minimal presence of macrophages (**Table S1**). A Masson’s trichrome staining highlighted interstitial collagen fiber deposition, suggesting fibrosis (**Figure 1D**), which were also confirmed as fibrosis in hematoxylin-eosin (HE) staining and under electron microscopy (**Figure 1E, 1F, 1G,** and **Figure S2A**). Both HE staining and electron microscopy revealed loss of myofibrillar bundles, interstitial oedema and necrosis (**Figure 1F, 1H, 1I, Figure S2B,** and **S2C**), which indicated myocytes degeneration. Notably, electron microscopy examination revealed lipofuscin granules accumulation in myocardial cells (**Figure 1J** and **Figure S2D**), the same kind of lipofuscin pigments was previously observed in sudden cardiac death (Kakimoto et al., 2019). Interestingly, we observed disordered myocardial ultrastructure in the biopsy tissue, especially in the mitochondria: a significant number of mitochondria were swollen and vacuolated, with distorted and broken cristae (**Figure 1K**, **1L,** and **Figure S2E**). Swollen and degenerating mitochondria were previously observed in autoimmune myocarditis in rat models (Skrzypiec-Spring et al., 2018; Skrzypiec-Spring et al., 2021). Unsurprisingly, we did not observe any SARS-CoV-2 virus in the endomyocardial biopsy sample, since the patient has test negative for almost a month. The immunohistochemical analysis, Masson’s trichrome staining, HE staining, and electron microscopic analysis aid in the diagnosis of myocarditis (Sagar et al., 2012). Throughout his hospitalization, Patient #1 received guideline-based care, including cardiac monitoring, oxygen therapy, and antiarrhythmic medications, leading to a stable discharge after ten days.

### Ultrastructural disorder in cardiomyocytes in a patient suffer chest tightness, palpitation and fatigue

As shown in **Table 1**, Patient #2 is a 60’s male who presented with Long COVID symptoms including chest tightness, palpitations, and fatigue persisting for one month prior to hospital admission. His medical history was notable for hypertension, with no other chronic cardiovascular conditions. Initial diagnostics revealed abnormal Q waves and ST segment depression in the III and avF leads on the ECG, and an Echo indicated impaired diastolic heart function. Laboratory tests showed mildly elevated troponin T (cTnT) levels. CAG found no evidence of coronary stenosis, but CMR imaging displayed delayed enhancement in the anterior septal muscle layer of the left ventricle’s basal and middle levels, suggestive of myocardial fibrosis.

**Table 1.** Summary data for 5 patients.

|  | Patient #1 | Patient #2 | Patient #3 | Patient #4 | Patient #5 |
| --- | --- | --- | --- | --- | --- |
| **Sex** | Male | Male | Male | Male | Male |
| **Age** | 26-30 | 66-70 | 41-45 | 65-70 | 31-35 |
| **Days between COVID-19 infection and myocardial biopsy** | 37 | 38 | 58 | 76 | 85 |
| **Diagnosis** | 1. Viral myocarditis<br>2. Sudden cardiac death<br>3. Ventricular fibrillation | 1. Viral myocarditis<br>2. Hypertension | 1. Viral myocarditis | 1. Viral myocarditis<br>2. Atrial fibrillation<br>3. Personal history of cerebral infarction | 1. Viral myocarditis |
| **Symptoms** | syncope | chest tightness<br>palpitation<br>fatigue | chest tightness<br>fever | chest tightness<br>dyspnea | chest pain |
| **ECG** | ventricular fibrillation | abnormal Q waves<br>ST segment depression | sinus tachycardia,<br>elevation of ST segments of II, III, avF, V5, V6 leads and abnormal Q waves of III, avF leads | frequent ventricular premature beats, frequent atrial premature beats, Atrial fibrillation<br>ST-segment depression | normal |
| **Cardiac biomarkers** | cTnT ↑<br>CK-MB ↑<br>Myoglobin ↑ | cTnT ↑ | cTnT ↑<br>Myoglobin ↑<br>CK-MB ↑<br>NT-pro BNP ↑ | cTnT ↑<br>NT-pro BNP ↑ | cTnT ↑<br>CK-MB ↑<br>Myoglobin ↑<br>NT-pro BNP ↑<br>CRP ↑ |
| **Echo** | no anomalies | decreased diastolic function of the heart | no anomalies | no anomalies | no anomalies |
| **CAG** | clean LAD, LCX and RCA arteries, Normal for EP study | no significant stenosis | 40% stenosis of LAD arteries | clean LAD, LCX and RCA arteries | clean LAD, LCX and RCA arteries |
| **CMR** | no anomalies | delayed enhancement of the anterior septal muscle layer at the basal and middle levels of the left ventricle, indicating myocardial fibrosis | myocardial edema in the left ventricular free wall and adjacent inferior wall | myocardial edema in the left ventricular septum, anterior wall, adjacent anterior septum, and free wall | no anomalies |
| **Pathological and immunohistochemical analysis** | interstitial collagen fiber deposits<br>IHC:<br>CD3 (-)<br>CD4 (-)<br>CD8 (-)<br>CD9 (-)<br>CD20 (-)<br>CD68 (weekly +)<br>Masson (+) | Slight cellular hypertrophy, interstitial collagen fiber deposits<br>IHC:<br>CD3 (-)<br>CD4 (-)<br>CD8 (-)<br>CD9 (-)<br>CD20 (-)<br>CD68 (weekly +)<br>Masson (+) | Slight cellular hypertrophy, IHC:<br>CD3 (-)<br>CD4 (-)<br>CD8 (-)<br>CD9 (-)<br>CD20 (-)<br>CD34 (+)<br>CD68 (weekly +)<br>Masson (+) | IHC:<br>CD3 (-)<br>CD4 (-)<br>CD8 (-)<br>CD9 (-)<br>CD20 (-)<br>CD34 (weekly +)<br>CD68 (-)<br>Masson (+) | Masson (+) |
| **Electron microscopy** | 1. local myofibrillar fibrosis<br>2. loss of integrity of myofibrillar bundles<br>3. Interstitial oedema and necrosis<br>4. lipofuscin granules accumulation<br>5. vacuolations in the mitochondria | 1. local myofibrillar fibrosis<br>2. loss of integrity of myofibrillar bundles<br>3. Interstitial oedema and necrosis<br>4. lipofuscin granules accumulation<br>5. vacuolations in the mitochondria | 1. local myofibrillar fibrosis<br>2. loss of integrity of myofibrillar bundles<br>3. Interstitial oedema and necrosis<br>4. lipofuscin granules accumulation<br>5. vacuolations in the mitochondria | 1. local myofibrillar fibrosis<br>2. loss of integrity of myofibrillar bundles<br>3. Interstitial oedema and necrosis<br>4. lipofuscin granules accumulation<br>5. vacuolations in the mitochondria | 1. local myofibrillar fibrosis<br>2. loss of integrity of myofibrillar bundles<br>3. Interstitial oedema and necrosis<br>4. lipofuscin granules accumulation<br>5. vacuolations in the mitochondria |

An endomyocardial biopsy performed 38 days post-infection revealed significant findings. Masson’s trichrome staining highlighted interstitial collagen fiber deposition (**Figure 2A** and **2B**). HE staining demonstrated interstitial collagen accumulation, disrupted myofibrillar bundle integrity, and interstitial infiltration by inflammatory cells (**Figure 2C** and **2D**). Immunohistochemical analysis showed a lack of reactivity for CD3, CD4, CD8, CD9, and CD20, with only a weak positive signal for interstitial CD68 (**Table 1**). Electron microscopy further detailed local myofibrillar fibrosis, fibroblast proliferation, muscle bundle edema, loss of myofibrillar integrity, presence of lipofuscin granules in cardiomyocytes, and significant mitochondrial vacuolations (**Figure 2E-2J**), mirroring the pathological changes observed in Patient #1 and reinforcing the diagnosis of myocarditis. Throughout his hospitalization, the patient underwent rigorous cardiac monitoring and was administered a comprehensive treatment regimen that included antiplatelet, antihypertensive, lipid-regulating, and myocardial remodeling-improving medications as secondary prevention measures. Following a stable 10-day hospitalization without any Major Adverse Cardiovascular Events (MACE), he was discharged, marking a critical step towards recovery.

**Figure 2.**
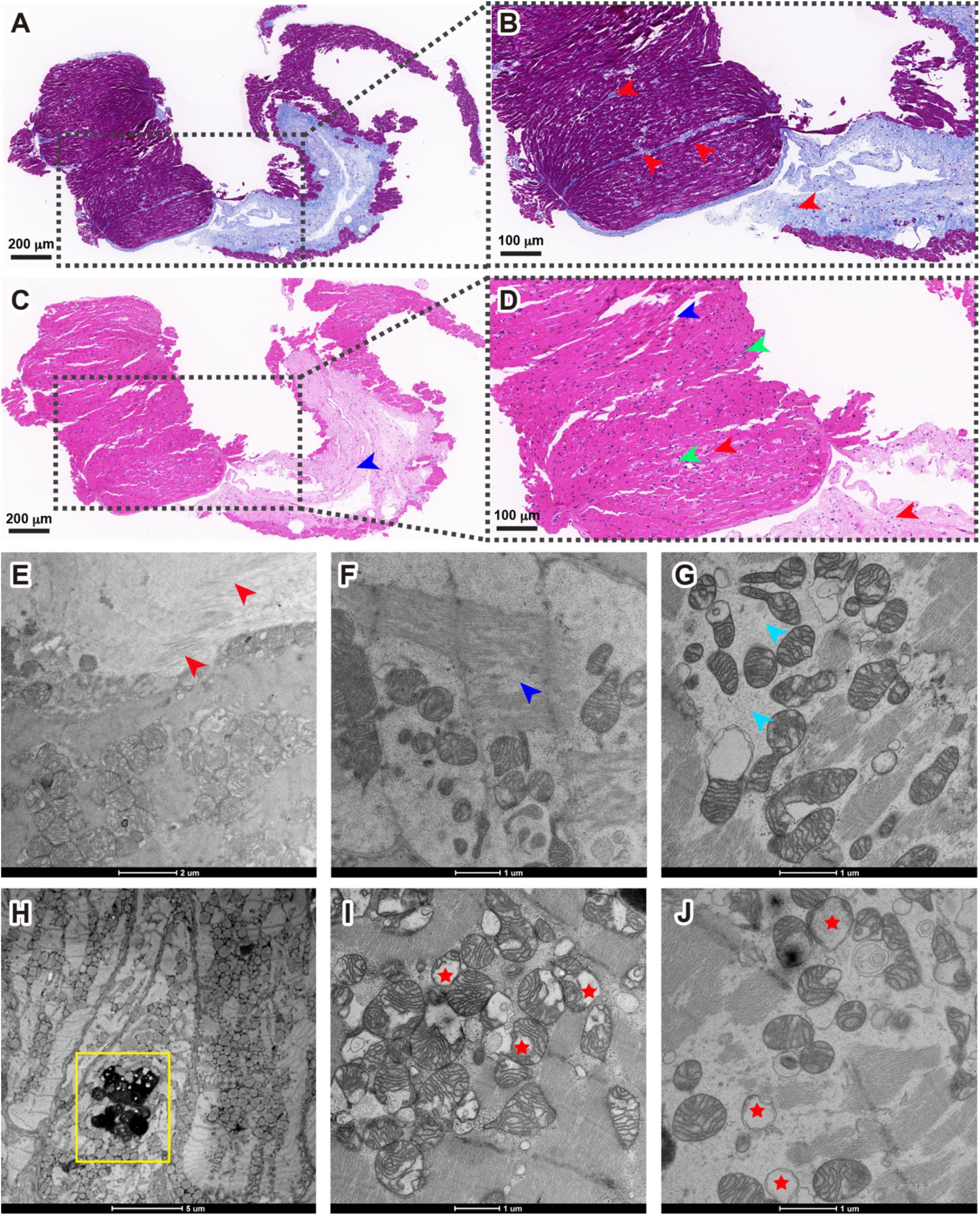
Spectrum of myocarditis in COVID-19 Patient #2. (A) Masson’s trichrome staining. (B) zoomed view displayed interstitial collagen fiber deposits (red arrowheads). (C) HE analysis shows an abnormal structure in the myocardium; (D) zoomed view, shows interstitial collagen fiber accumulation (red arrowheads), loss of integrity of myofibrillar bundles (blue arrowheads), and myofibrillar interstitial infiltration with inflammatory cells is also observed (green arrowheads). (E) electron micrographs of the biopsy sample from the patient, showing local myofibrillar fibrosis and proliferation of fibroblasts (red arrowheads); (F) loss of integrity of myofibrillar bundles (blue arrowheads); (G) necrosis of myofibrillar bundles (cyan arrowheads); (H) lipofuscin granules in the cardiomyocytes (yellow frames); (I) and (J) a significant amount of vacuolations in the mitochondria (red stars).

### Ultrastructural disorder in cardiomyocytes in three patients diagnosed with myocarditis

Patient #3 is a 40’s male who recovered from COVID-19 two months prior to admission. This patient does not have any chronic diseases but presented with symptoms of fever and chest tightness. Pathogenetic tests, including SARS-CoV-2 and coxsackievirus, were shown to be negative. ECG suggested sinus tachycardia, elevated ST segments in leads II, III, avF, V5, and V6, as well as abnormal Q waves in III and avF leads. Cardiac enzyme markers, including cTnT, myoglobin, creatine kinase isoenzyme MB (CK-MB), and N-terminal pro-brain natriuretic peptide (NT-pro BNP), were all significantly elevated, indicating myocardial injury. CMR confirmed acute myocarditis, primarily affecting the left ventricular free wall and the proximal region of the inferior wall. A small amount of pericardial effusion and impaired left ventricular systolic function were also observed. Other imaging tests, including chest CT, Echo, and CAG, did not reveal significant abnormalities.

Patient #4 is a 70’s male who recovered from COVID-19 two and a half months before being admitted to our hospital. He does not have any chronic diseases but presented with symptoms of chest tightness and dyspnea. ECG revealed sinus rhythm, frequent ventricular premature beats, frequent atrial premature beats, atrial fibrillation, and ST-segment depression. Cardiac enzyme NT-pro BNP levels were elevated. CMR indicated myocardial edema in the left ventricular septum, anterior wall, adjacent anterior septum, and free wall, suggesting myocarditis. Additional imaging tests revealed no significant abnormalities.

Patient #5 is a 30’s male who recovered from COVID-19 three months prior to admission. He experienced chest pain, along with elevated cardiac enzyme markers and an increased C-reactive protein (CRP) level. Imaging tests, including ECG, CMR, Echo, chest CT, and CAG, did not indicate significant changes, suggesting a subclinical presentation of myocarditis.

All three patients were diagnosed with myocarditis based on clinical presentation, elevated cardiac markers, and CMR findings. Endomyocardial biopsies conducted at various intervals post-COVID-19 infection revealed interstitial collagen deposits, lipofuscin granules accumulation, loss of integrity and necrosis of myofibrillar bundles, and notably, mitochondrial ultrastructural disorders similar to patients #1 and #2, reinforcing the diagnosis of myocarditis (**Figure 3** and **Figure S3**).

**Figure 3.**
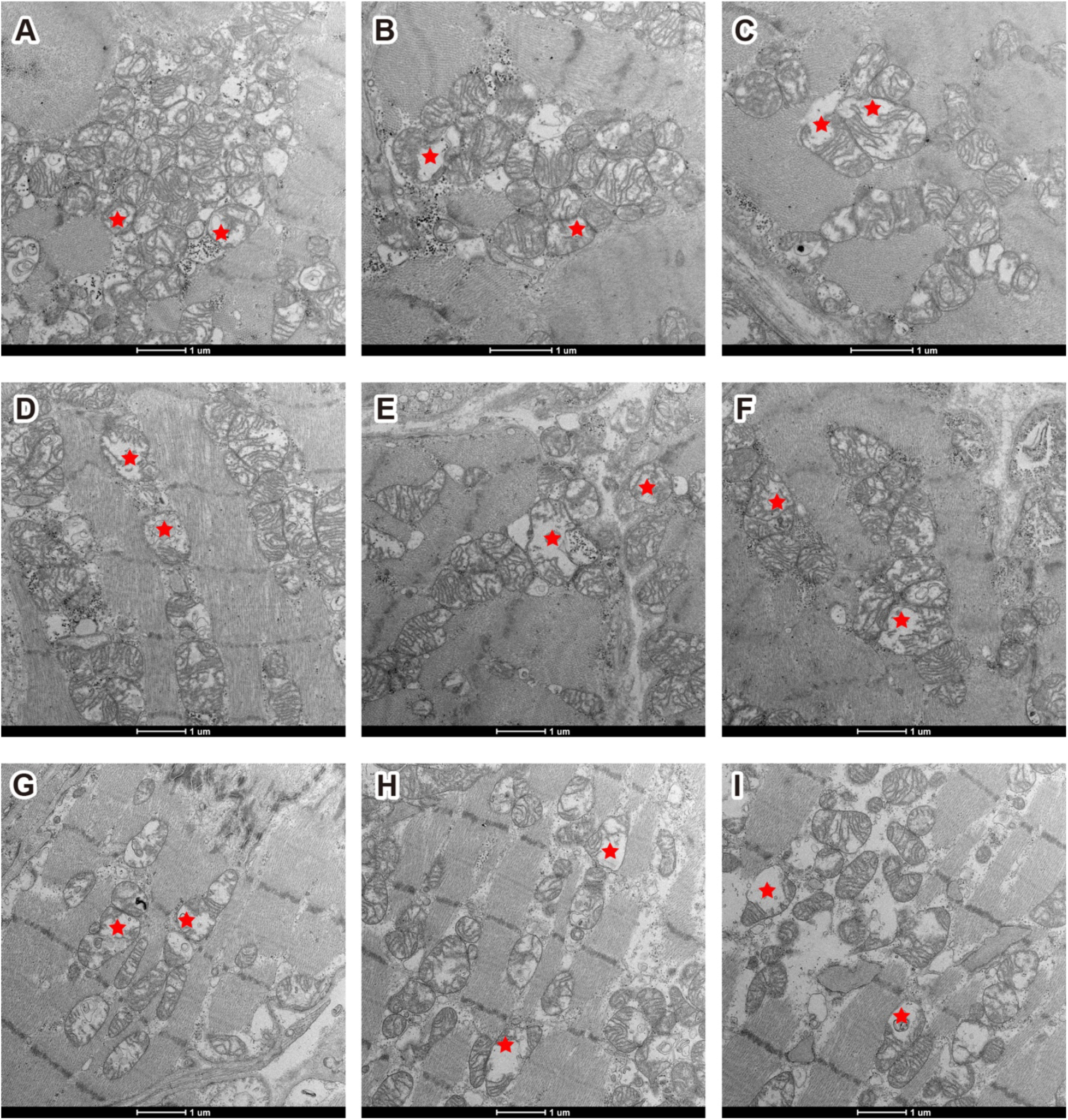
Mitochondria disorders in COVID-19 patient #3, #4, and #5. (A)-(C) patient #3. (D)-(F) patient #4. (G)-(I), patient #5.

### Qualitative and quantitative analysis of mitochondria disorder in 2D images and 3D volumes

We conducted a qualitative analysis of mitochondria disorder rate in the myocardial biopsy samples obtained from five patients. Forty cardiomyocytes from each patient were randomly picked, with approximately 80-100 mitochondria in each cardiomyocyte were selected for analysis. We classified them into two groups: healthy or disordered mitochondria, and the percentage of the disordered mitochondria rate was calculated for each cardiomyocyte. As shown in **Figure 4A**, the proportions of disordered mitochondria relative to the total number of mitochondria are approximately 40-60% for all five patients. This suggests that there is no significant difference in the extent of mitochondrial damage between what we speculated as longer recovery periods (e.g., 76 days and 85 days) and shorter recovery periods (e.g., 37 days and 38 days). It should be noted that the results in **Figure 4A** did not account for various influencing factors such as patient age, pre-existing health conditions before contracting COVID-19, and the presence of any underlying diseases.

**Figure 4.**
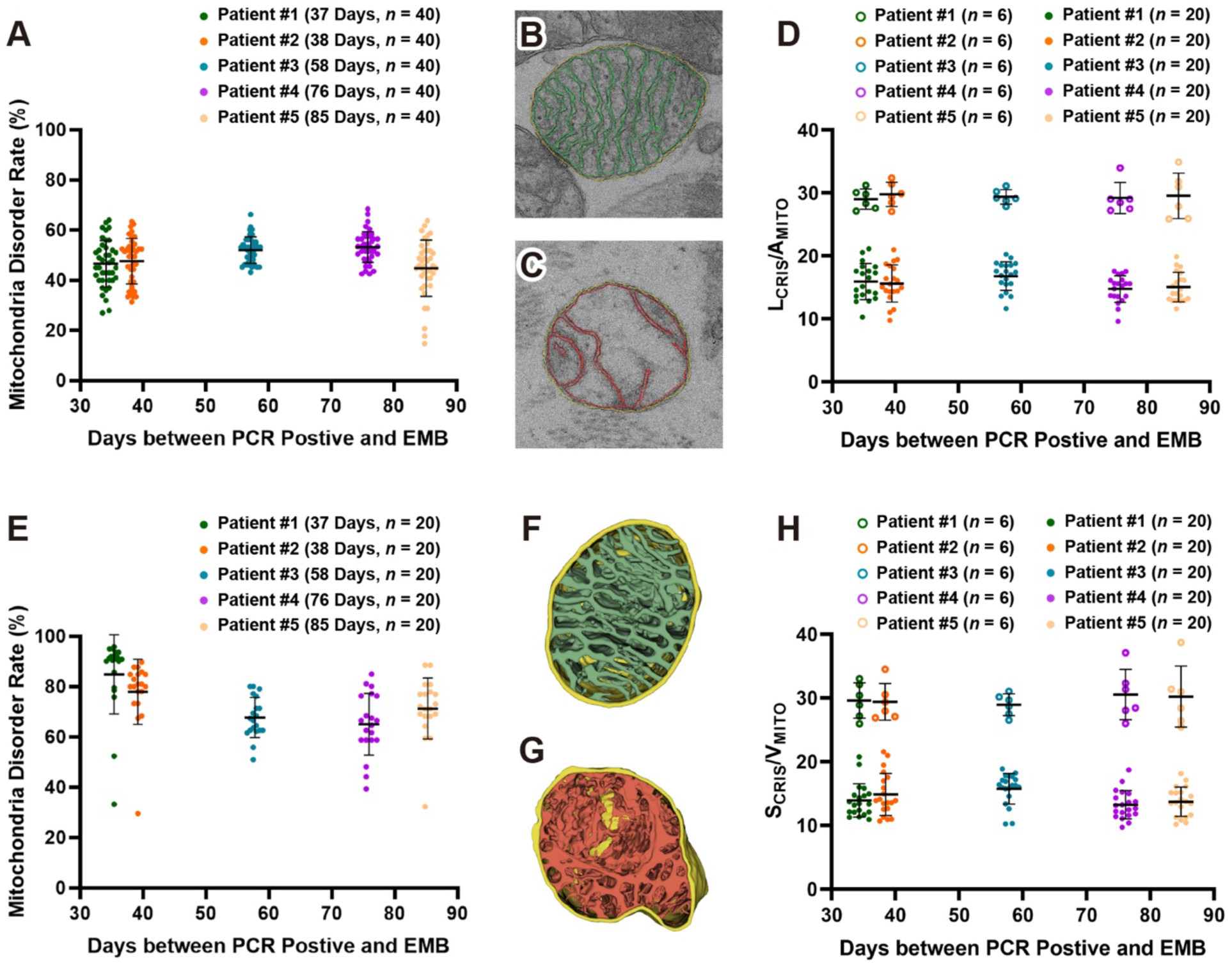
Qualitative and quantitative analysis of mitochondria disorder in 2D images and 3D volumes. (A) analysis of mitochondria disorder rate in 40 cardiomyocytes from each patient, each point represents an averaged percentile of disordered mitochondria from total 80-100 mitochondria in each cardiomyocyte. (B) and (C) 2D segmentation of outer membrane, inner membrane, and cristae in a healthy and disordered mitochondrion, respectively. (D) analysis of mitochondria disorder by comparison the ratio of entire cristae length to the area of the mitochondrial cross section (L_CRIS_/A_MITO_). Healthy mitochondria like the one shown in Panel B have an averaged ratio around 29 μm^-1^ (open circles in top portion of Panel D). In contrast, disordered mitochondria like the one shown in Panel C have a lower ratio (filled circles in lower portion of Panel D). (E) analysis of mitochondria disorder rate in 20 tissue blocks from each patient, each point represents an averaged percentile of disordered mitochondria from total 80-100 mitochondria in each block. (F) and (G) 3D segmentation of outer membrane, inner membrane, and cristae in a healthy and disordered mitochondrion, respectively. (H) analysis of mitochondria disorder by comparison the ratio of surface area of the entire cristae contained within a mitochondrial volume (S_CRIS_/V_MITO_). Healthy mitochondria like the one shown in Panel F have an averaged ratio around 27 μm^-1^ (open circles in top portion of Panel H), disordered mitochondria like the one shown in Panel G have lower ratios (filled circles in lower portion of Panel H).

Next, we performed segmentation of the outer mitochondrial membrane (OMM), inner mitochondrial membrane (IMM), and cristae in 2D electron microscope images. **Figures 4B** and **4C** display segmentation marks for a healthy mitochondrion and a disordered mitochondrion, respectively. We measured the length of the entire cristae (the inner boundary membrane plus the cristae membranes) and calculated the ratio of entire cristae length to the area of the mitochondrial cross section (L_CRIS_/A_MITO_), termed “crista density”, measured in 2D images (Adams et al., 2023). This ratio quantifies the degree of damage to the mitochondrial cristae structure. As shown in **Figure 4D**, the more severe the structural damage to the mitochondria, the more pronounced the cristae loss, resulting in a diminished length of the cristae membrane and a reduced ratio of L_CRIS_/A_MITO_. Our data showed that this ratio in healthy human mitochondria is around 29 μm^-1^, as depicted by open circles in top portion in **Figure 4D**. In contrast, the ratio dramatically decreased in the disordered mitochondria, and in the worst-case scenario, almost cristae membranes had vanished in the vacuolar mitochondria, and the ratio drops close to 10 μm^-1^ (filled circles in lower portion in **Figure 4D**). Previous studies have reported that L_CRIS_/A_MITO_ ratios varying from 34, 37, to 41 μm^-1^ in rat myocardial cells (Adams *et al*., 2023; El’darov et al., 2015; Mall et al., 1980), which are slightly higher than those we measured in human cardiomyocytes.

We further analysis the mitochondria disorder in 3D volumes by focused ion beam scanning electron microscopy (FIB-SEM)(Kizilyaprak et al., 2019), one of the 3D volume electron microscopy techniques that is promised to have an outsized impact on science in the coming years (Eisenstein, 2023; Peddie et al., 2022). As shown in **Movie S1**, a tissue fragment from Patient #1 was analyzed by FIB-SEM, a gallium FIB was used to shave 10nm thin section from the tissue fragment followed by SEM imaging (pixel size 5nm, in X-Y panel), 930 serial images were binned (convert X-Y pixel size to 10nm to make 10nm isotropic resolution in X-Y-Z direction), aligned, cropped (remove misaligned edges), and stacked to form a 3D volume measuring 16μm×12μm×9.3μm in X-Y-Z dimensions.

Next, OMM, IMM, and cristae membrane of each mitochondrion were segmented using a deep learning-based MitoStructSeg platform (Wang et al., 2024), which employs an Adaptive Multi-Domain Mitochondrial Segmentation (AMM-Seg) model specifically developed to accurately segment OMM, IMM, and cristae membranes of mitochondria. As shown in **Movie S2**, a volume block was selected from a FIB-SEM raw dataset (4μm×4μm, 40 serial images) and displayed in two-channel: raw images and AI-based segmentation images. The raw image is initially submitted to AMM-Seg, where it undergoes rigorous data preprocessing to yield an augmented image and a textured image. Subsequently, the encoder precisely extracts the distinctive features from both the augmented and textured images. The adaptive fusion module then ingeniously integrates these two feature sets to create an enhanced and comprehensive feature representation. Finally, the decoder utilizes the enhanced representation to generate the mitochondrial segmentation results, including OMM, IMM and cristae membranes.

**Movie S3** displayed a tissue block that enriched with mitochondria, measuring 4μm×4μm×9μm, with all mitochondria were automatically segmented in the block. We classified them into two groups: healthy (shown in green, a selection criterion was that no cristae damage was observed in any section of the 3D volume) and disordered (shown in red). The percentage of the disordered mitochondria rate was calculated. Twenty blocks from each of the five patients were analyzed, with approximately 80-100 mitochondria in each block were selected for analysis. **Figure 4E** indicated that the proportion of disordered mitochondria rate is approximately 60-90% for all five patients, more severe than those analyzed in 2D images (**Figure 4A**). This indicated that mitochondria disorder rates measured in 3D volume are more accurate than those measured in 2D images.

Figure 4F and **4G** illustrated 3D visualization of OMM, IMM, and cristae in a healthy mitochondrion and a disordered mitochondrion, respectively. We quantified crista density in 3D volumes by measure the surface area of the entire cristae contained within a mitochondrial volume (S_CRIS_/V_MITO_). Figure 4H indicated that ratios in healthy human mitochondria are around 28 μm^-1^ (depicted as open circles in top portion in Figure 4H). In contrast, the ratio decreased in the disordered mitochondria (filled circles in lower portion in Figure 4H). The crista density ratios quantified in 3D volumes as S_CRIS_/V_MITO_ are consistent with L_CRIS_/A_MITO_ ratios quantified in 2D images (Figure 4D).

### Similar mitochondria disorganization found in SARS-CoV-2 Omicron infected mice

We hypothesized the ultrastructural disorders observed in the cardiomyocytes of patients were caused by SARS-CoV-2 infection. Due to limited resource for endomyocardial biopsy samples and the lack of healthy controls, we investigated these ultrastructural disorders using mice infected with SARS-CoV-2. According to a recent assessment of the diversity of Omicron sublineages and the epidemiologic features of the autumn/winter 2022 COVID-19 wave in Shanghai, BA.5.2 and BQ.1 are the two dominating sublineages (Lu et al., 2023). In our animal study, BALB/c mice were challenged with Omicron BA.5.2 and BQ.1 sublineages, heart tissues were collected 7 days post infection (dpi, convalescence stage for mouse infection).

We observed similar disorganization in mitochondria and in myofibrillar bundles in heart tissues of both BA.5.2 and BQ.1 infected mice (4 mice in each group, Figure 5A and **5B**). In contrast, the control mice showed normal mitochondrial ultrastructure (Figure 5C). The structural alterations in mitochondria are consistent with findings in murine models of viral myocarditis (Helluy et al., 2017; Zhang et al., 2020). However, we did not observe any collagen fiber or lipofuscin granule in the mice heart.

**Figure 5.**
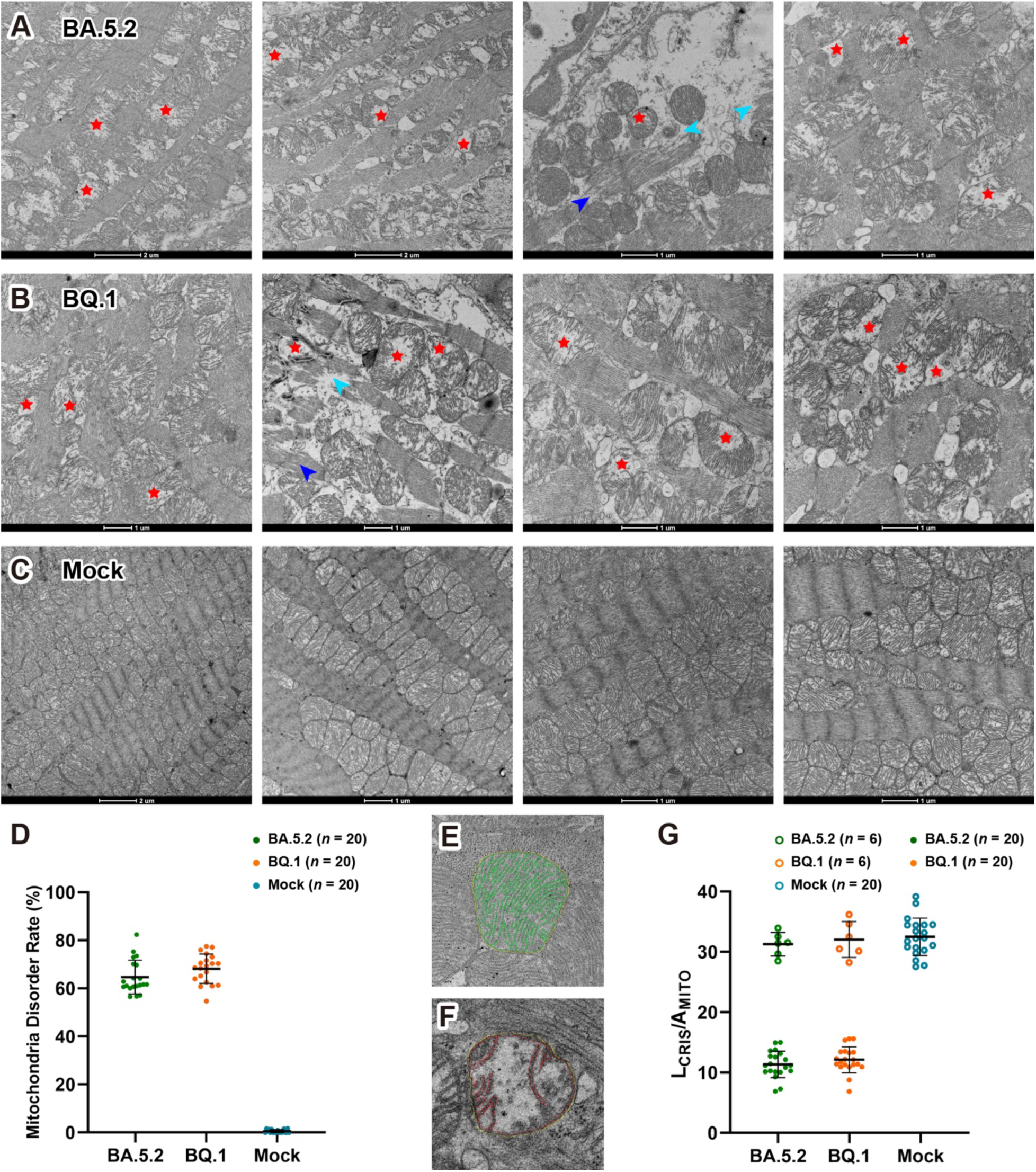
Mitochondria disorganization in SARS-CoV-2 infected mice. (A) and (B), electron micrographs of heart tissue from BA.5.2 and BQ.1 infected mice, showing similar disorder (red stars) in the mitochondria and myofibrillar bundles (blue and cyan arrowheads), as observed in the biopsy tissue from the patient. (C) electron micrographs of heart tissue from control group, showing normal mitochondria ultrastructure. (D) analysis of mitochondria disorder rate in 20 cardiomyocytes from each mouse, each point represents an averaged percentile of disordered mitochondria from total 80-100 mitochondria in each cardiomyocyte. (E) and (F) 2D segmentation of outer membrane, inner membrane, and cristae in a healthy and disordered mitochondrion, respectively. (G) analysis of mitochondria disorder by comparison the ratio of entire cristae length to the area of the mitochondrial cross section (L_CRIS_/A_MITO_). Healthy mitochondria like the one shown in Panel E have an averaged ratio around 30 μm^-1^ (open circles in top portion of Panel G). In contrast, disordered mitochondria like the one shown in Panel F have a lower ratio (filled circles in lower portion of Panel G).

Furthermore, we analyzed the mitochondria disorder rate and cristae density ratio in 2D images, Figures 5D exhibites that the mitochondria disordered rates are round 60-75% in both BA.5.2 and BQ.1 infected mice, whereas no disordered mitochondria were found in the mock group. Figures 5E**-5G** indicate that cristae density L_CRIS_/A_MITO_ ratio in healthy human mitochondria is around 32 μm^-1^. In contrast, the rates in disordered mitochondria, markedly decreased to 9-15 μm^-1^, indicating a significant loss of cristae ultrastructure in both BA.5.2 and BQ.1 infected cardiomyocytes.

### Proteomic profiling of SARS-CoV-2 infected mouse heart tissues

We performed quantitative mass spectrometry analysis of heart tissues from BA.5.2 and BQ.1 infected mice, as well as from a mock group (*n*=4, Figure 6). Specifically, when comparing the BA.5.2 group to the mock group, 36 differentially expressed proteins (DEPs) were significantly upregulated, and 41 DEPs were significantly downregulated (Figures 6A and **6B**). When comparing the BQ.1 group to the mock group, 109 DEPs were significantly upregulated, and 10 DEPs were significantly downregulated (Figures 6C and **6D**). To explore the potential functions of these differentially expressed proteins, we conducted Kyoto Encyclopedia of Genes and Genomes (KEGG) pathway enrichment and found functional overlaps between BA.5.2 and BQ.1. The enriched KEGG pathways of DEPs in were involved in oxidative phosphorylation, cardiac muscle contraction, and mitophagy (Figures 6E and **6F**).

**Figure 6.**
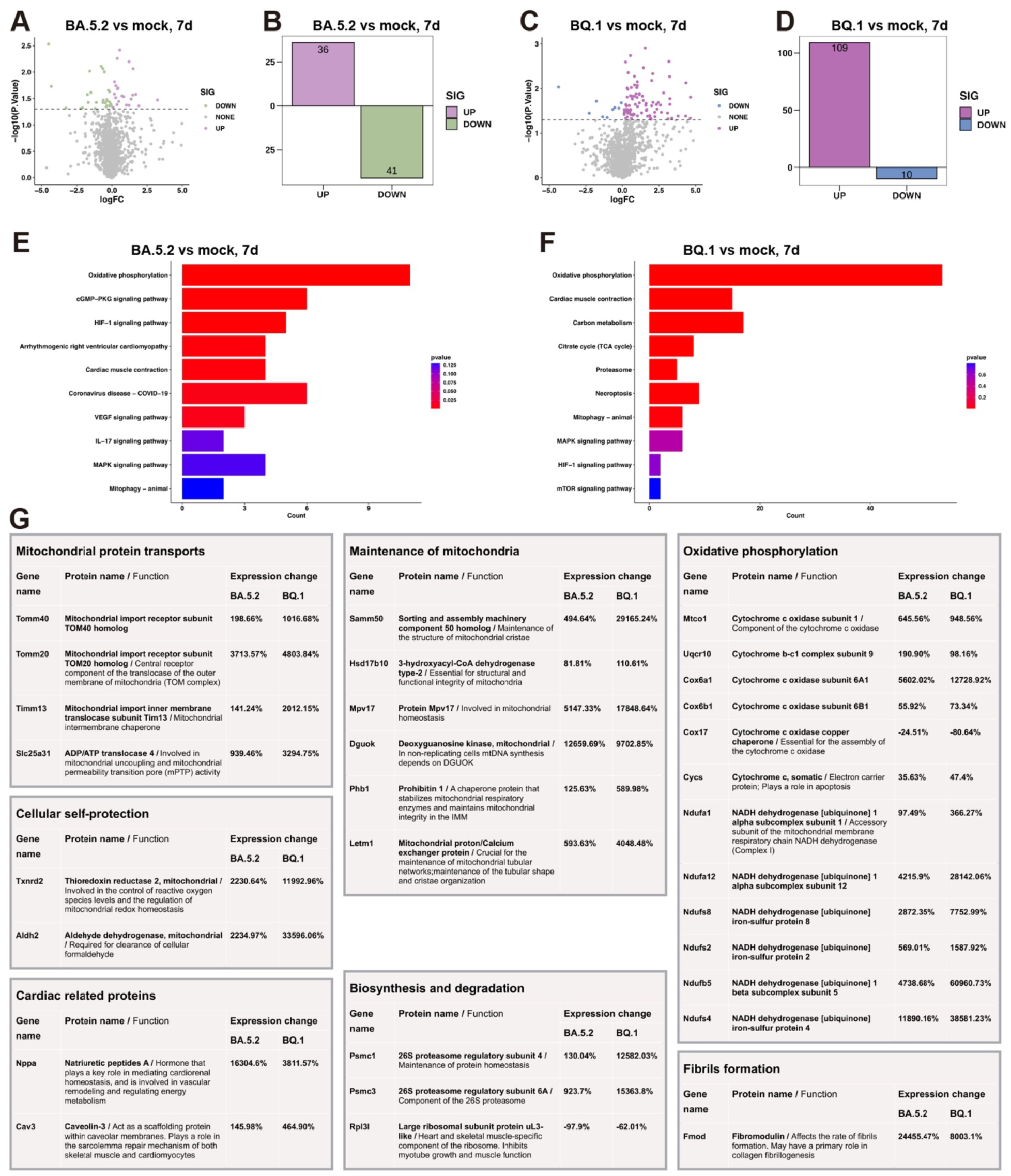
Proteomic profiling of SARS-CoV-2 infected mouse heart tissues. (A) and (C) volcano plots representing differentially abundant proteins in BA.5.2 and BQ.1 infected mice heart tissue compared with mock group (n = 4). Green and blue dots represent proteins with a log2 fold change < –0.5 and a q value < 0.05. Plum and purple dots represent proteins with a log2 fold change > 0.5 and a q value < 0.05. Gray dot line indicates q value = 0.05. Only proteins with >2 identified unique peptides were considered. (B) and (D), numeric quantification of colored data points in A and C. (E) and (F) KEGG pathway enrichment analysis of significantly changed proteins. (G) groups of proteins that upregulated or downregulated in BA.5.2 and BQ.1 infected mice heart, percent changes in protein level are indicated for each protein.

Further functional enrichment analysis indicated that the most significantly changed genes and proteins in BA.5.2 and BQ.1 infected heart were related to mitochondria (Figure 6G), including mitochondrial protein transports (mitochondrial import receptors TOM40 and TOM20, mitochondrial import inner membrane translocase TIM13, and Slc25a31 ADP/ATP translocase); maintenance of mitochondria (Sorting and assembly machinery component Samm50, dehydrogenase Hsd17b10, Protein Mpv17, Deoxyguanosine kinase Dgouk, Prohibitin 1, and mitochondria protin/calcium exchanger Letm1); oxidative phosphorylation (cytochrome c oxidases, cytochrome bc1 complex, and NADH dehydrogenases); and cellular self-protection (mitochondrial thioredoxin reductase 2 and mitochondrial aldehyde dehydrogenase). Most proteins related to mitochondria are upregulated, likely compensating for protein lost, damage repair, and the generation of new mitochondria.

*Mitochondrial protein transports* (1). TOM40: Most mitochondrial proteins are synthesized as precursors in the cytosol and imported by the translocase of the outer mitochondrial membrane (TOM). The β-barrel protein TOM40 forms the protein-conducting channel of the TOM complex and acts as the central porin of TOM complex to import of precursor proteins into the inter-OMM-IMM space (Dolezal et al., 2006). Consequently, deficits in TOM40 adversely affect preprotein import, disrupting mitochondrial homeostasis and cell viability (Baker et al., 1990) and arresting animal growth (Billing et al., 2011). On the other hand, over-expression of TOM40 is associated with elevated mitochondrial membrane potentials, respiratory rates, cellular ATP levels, suggesting that increased protein level of TOM40 may be protective of mitochondrial function (Zeitlow et al., 2017). (2). TOM20 is an OMM protein that functions as a component of the TOM complex. The primary role of TOM20 is receptor recognition, targeting mitochondrial precursor proteins and guiding them to TOM40 for protein translocation (Endo and Kohda, 2002). Tom20 depletion is known to induce a grossly altered morphology of mitochondria, Tom20-deficient fungi contain mitochondria that are highly deficient in cristae ultrastructure (Harkness et al., 1994). The lack of cristae in TOM20-deficient mitochondria is similar to what we observed in human and mouse cardiomyocytes post SARS-CoV-2 infection. It remains unclear if SARS-CoV-2 affects TOM40 and TOM20, nevertheless, a recent study revealed that the ORF9b protein of both SARS-CoV and SARS-CoV-2 physically interacts with TOM70, a partner of TOM20 that also recognizes and transfers mitochondrial preproteins to TOM40, and that the expression level of TOM70 was found decreased during SARS-CoV-2 infection (Gordon et al., 2020). (3). TIM13 is a member of translocase of the inner mitochondrial membrane (TIM) family of chaperones, TIM13 functions in the inter-OMM-IMM space to shield unfolded, hydrophobic membrane proteins and maintain an import-competent state for downstream translocation (Beverly et al., 2008; Koehler, 2004). Additionally, a recent study demonstrated that TIM13 is closely related to fibrosis in liver disease, silencing TIM13 significantly reduced the expression of profibrogenic genes and apoptosis related genes, although the underlying mechanism has yet to be fully elucidated (Liao et al., 2023). (4). Slc25a31 is a member of the ADP/ATP carrier family of proteins that exchange cytosolic ADP for matrix ATP in the mitochondria. Over-expressing of Slc25a31 has been shown to display an anti-apoptotic phenotype (Gallerne et al., 2010).

*Maintenance of mitochondria* (1). Samm50 is the core component of the sorting and assembly machinery (SAM) that plays a key role in the OMM, and it has been confirmed that Samm50 promotes the biogenesis of TOM40 by forming a SAM-TOM super-complex, and Samm50 deficiency inhibits the assemble of TOM40 (Humphries et al., 2005; Qiu et al., 2013). This disruption can lead to impaired mitochondrial biogenesis and maintenance, resulting in decreased mitochondrial function and altered energy metabolism. Studies have shown that reduced expression of Samm50 leads to the development of myocardial hypertrophy and associated mitochondrial dysfunctions, including impaired mitophagy and increased oxidative stress in mouse ventricular cardiomyocytes (Xu et al., 2021). Moreover, Samm50 physically interacts with Mitofilin and CHCHD6, two core proteins of the mitochondrial contact site and cristae organizing system (MICOS) complex in the junction of IMM and cristae, Samm50 is indispensable for cristae structure stability and the proper assembly of the mitochondrial respiratory chain complexes (Ding et al., 2015). Depletion of Samm50 leads to changes in mitochondrial shape and the loss of mitochondrial cristae (Ott et al., 2015; Ott et al., 2012). (2). Hsd17b10 encodes 3-hydroxyacyl-CoA dehydrogenase type-2, a member of the short chain dehydrogenase/reductase superfamily. This mitochondrial protein is involved in pathways of fatty acid, branched chain amino acid and steroid metabolism and has been reported to be associated with mitochondrial toxicity in neurodegenerative diseases (Lustbader et al., 2004). Loss of Hsd17b10 function mediated by gene mutation, knock-down, or knock-out causes mitochondrial dysfunction and apoptotic cell death. Moreover, EM analysis of mitochondria from *Xenopus* embryos, mouse brain, and human fibroblasts showed that Hsd17b10 protein is required for structural and functional integrity of mitochondria (Rauschenberger et al., 2010). (3). MPV17 is an IMM protein and its deficiency can cause mitochondrial DNA (mtDNA) depletion, increase reactive oxygen species (ROS), and promote mitochondrial apoptosis (Dalla Rosa et al., 2016; Zwacka et al., 1994). Moreover, Electron microscopy examination observed broken or disappearance of cristae in MPV17 knock-out zebrafish and mice, and in Sym1 (yeast ortholog of MPV17) ablation yeast (Bian et al., 2021; Dallabona et al., 2010; Viscomi et al., 2009). (4). *DGUOK* gene encodes mitochondrial deoxyguanosine kinase, which mediates phosphorylation of purine deoxyribonucleosides in the matrix. DGUOK is crucial for the maintenance and replication of mtDNA (Petrakis et al., 1999). DGUOK deficiency leads to mtDNA depletion syndrome (MDS), which results in reduced mtDNA content and decreased activity of the mtDNA-encoded respiratory chain complexes I, III, IV, and V (Taanman et al., 1997). The depletion impairs the mitochondrial respiratory chain function, leading to decreased ATP production and increased production of ROS. The resulting oxidative stress further damages mitochondrial components, exacerbating the dysfunction. DGUOK-related MDS is characterized by structural abnormalities in mitochondria, such as loss of cristae integrity and swollen mitochondria (Lee et al., 2009; Mandel et al., 2001). (5). Prohibitin 1 (PHB1), along with its homolog PHB2, forms a large complex in the IMM that is essential for maintaining mitochondrial integrity and functionality, Prohibitins are involved in regulating mitochondrial dynamics, including processes of fusion and fission which are crucial for mitochondrial adaptation under stress conditions; Prohibitins function as chaperone proteins that stabilizes mitochondrial respiratory enzymes and maintains mitochondrial integrity in the IMM and cristae (Signorile et al., 2019). When Prohibitins are deficient, there is a disruption in these dynamics, leading to impaired mitochondrial division and fusion, affecting the mitochondrial membrane potential and results in an aberrant cristae morphogenesis (Merkwirth et al., 2008). Over-expression of Prohibitin 1 in cardiomyocytes has been shown to protect cells from oxidative stress-induced mitochondrial membrane permeability, inhibited release of cytochrome c, and suppressed mitochondrial apoptosis (Liu et al., 2009). Additionally, mitochondrial shape and ultrastructure are affected by the lipid composition of mitochondria. Prohibitins affect the maturation of cardiolipin, a phospholipid of the mitochondrial inner membrane that plays a role in mitochondrial fusion (Richter-Dennerlein et al., 2014). (6). Letm1 is a mitochondria protin/calcium exchanger localized to the IMM (Jiang et al., 2009). Letm1 maintains the mitochondrial tubular shapes and is required for normal mitochondrial morphology and cellular viability, LETM1 downregulation cause mitochondrial swelling and cristae disorganization, formation of the respiratory chain complexes are also impaired by LETM1 knockdown (Tamai et al., 2008).

*Oxidative phosphorylation* Multiple genes and proteins involve in cytochrome c oxidase, cytochrome bc1 complex, and NADH dehydrogenas. NADH dehydrogenas known as Complex I, Cytochrome bc1 complex as Complex III, and cytochrome c oxidase as Complex IV of the mitochondrial electron transport chain. They are the enzymes of the mitochondria electron transport chain and play crucial roles in cellular energy production, the process is essential for maintaining the proton gradient that drives ATP synthesis. Impairment of NADH dehydrogenas, cytochrome bc1 complex, and cytochrome c oxidase activities disrupt the electron transport chain, leading to reduced ATP production, increased ROS production, and impaired cellular respiration. These changes result in various structural and functional abnormalities in mitochondria, including altered mitochondrial morphology, increased mitochondrial fragmentation, and swelling (Vercellino and Sazanov, 2022).

*Cellular self-protection* (1). Gene *Txnrd2* encodes thioredoxin reductase 2, a selenocysteine-containing enzyme essential for mitochondrial oxygen radical scavenging. Thioredoxin reductase 2 is crucial for maintaining redox homeostasis within mitochondria by reducing oxidative stress (Arnér and Holmgren, 2000). Cardiac-specific deletion of Txnrd2 in mice results in dilated cardiomyopathy, the lack of Txnrd2 results in increased oxidative damage, disrupted mitochondrial dynamics, and impaired energy production, leading to structural alterations such as mitochondrial fragmentation and loss of cristae integrity (Conrad et al., 2004). (2). Gene *Aldh2* encodes aldehyde dehydrogenase 2, an enzyme predominantly found in the mitochondrial matrix, crucial for detoxifying aldehydes generated during oxidative stress. ALDH2 deficiency leads to increased accumulation of toxic aldehydes, which impaired mitochondrial respiration, reduced ATP production, and increased ROS generation, leading to oxidative damage to mitochondrial DNA, proteins, and lipids (Wu and Ren, 2019). The lack of ALDH2 exacerbates mitochondrial structural abnormalities, including swelling and loss of cristae integrity (Kuroda et al., 2017).

Other proteins involved in fibrils formation (fibromodulin), biosynthesis and degradation (26S proteasomal subunits), as well as cardiac related proteins (natriuretic peptides A and caveolin-3), were upregulated in BA.5.2 and BQ.1 infected mice heart. One ribosomal subunit protein, Rpl3l, which inhibits myotube growth and muscle function in skeletal and cardiac muscle (Chaillou et al., 2016), was down-regulated.

## DISCUSSION

The global COVID-19 pandemic has presented clinicians with the daunting task of diagnosing and treating patients with non-specific cardiovascular symptoms, complicated by ambiguous CMR findings. Myocarditis, characterized as an inflammatory disease of the myocardium, requires immunological and histological confirmation for diagnosis, and endomyocardial biopsy serves to gain certainty about the diagnosis and identify the potential cause of the disease. Despite Patient #1 lacking typical viral myocarditis symptoms such as chest pain and dyspnea, elevated cardiac troponin levels prompted further investigation. Following the “2022 Expert Consensus Decision Pathway on Cardiovascular Sequelae of COVID-19 in Adults: Myocarditis and Other Myocardial Involvement, Post-Acute Sequelae of SARS-CoV-2 Infection, and Return to Play” by the American College of Cardiology (Gluckman et al., 2022), we proceeded with endomyocardial biopsy, aligning with international expert consensus that published in 2021 by the Heart Failure Association of the European Society of Cardiology, the Heart Failure Society of America, and the Japanese Heart Failure Society, in which endomyocardial biopsy is recommended in diagnostic assessment of select patients with atypical myocarditis (Seferović et al., 2021). In this study, immunohistochemical analysis, Masson’s trichrome staining, HE staining, and electron microscopy exam were performed on the endomyocardial biopsy samples, providing precise evidences for the diagnosis of myocarditis. We encourage clinicians to consider endomyocardial biopsy following the above expert consensuses, particularly in diagnosis of long-COVID associated cardiovascular outcomes.

The presence of lipofuscin granules in cardiomyocytes, indicative of cellular damage and aging, has been historically associated with myocardial hypertrophy, heart failure, and sudden cardiac death (Kajihara et al., 1973; Kakimoto *et al*., 2019). These granules result from the oxidation and polymerization of protein and lipid residues, often accumulating when mitochondria suffer structural damage, underscoring their significance as markers of cellular distress (Malkoff and Strehler, 1963). When mitochondria undergo structural damage, lysosomes, which are normally responsible for mitochondrial turnover, accumulate lipofuscin granules (Brunk and Terman, 2002). Our findings of mitochondrial disorganization and lipofuscin accumulation align with these observations, suggesting that SARS-CoV-2 infection contributes significantly to mitochondrial damage and accumulation of lipofuscin granules in the cardiomyocytes.

We have observed an interesting phenomenon in the mitochondria, the swollen and vacuolated mitochondria, distorted and broken cristae, all indicated severe damage to this important cellular organelle. This damage is highly likely caused by the SARS-CoV-2 Infection, as we have found an almost identical mitochondria disorganization in the mice that were infected with SARS-CoV-2. Our findings are consistent with a recent study in which mitochondrial gene expression was analyzed in autopsy tissues from patients with COVID-19, and core mitochondrial gene expression were suppressed in the hearts, chronically impaired mitochondrial function, and led to severe COVID-19 pathology in cardiovascular diseases (Guarnieri et al., 2023).

Mitochondrial damage has profound implications for cellular respiration, ATP production, and metabolism, potentially precipitating cardiovascular, neurodegenerative, and metabolic disorders. Notably, our observations indicate persistent mitochondrial damage three months post-COVID-19 recovery, challenging existing recommendations on exercise cessation post-infection. The ACC’s Sports and Exercise Cardiology Council’s recommendation for a minimum two-week cessation of exercise for mildly or moderately symptomatic COVID-19 athletes may need reevaluation (Phelan et al., 2020), considering our findings and prior guidance advocating for 3-6 months of exercise abstinence for individuals with clinical myocarditis (Maron et al., 2015).

This study’s limitation is its reliance on five patients’ data, underscoring the need for larger-scale studies to validate these findings and further elucidate the long-term cardiovascular impacts of COVID-19. Our research highlights the critical need for extended recovery periods before resuming exercise or strenuous activities post-COVID-19, based on the significant mitochondrial damage observed in cardiomyocytes.

## STAR METHODS

### Clinical study

Clinical study was approved by the Human Ethics Committee of Shanghai Tenth People’s Hospital, Tongji University School of Medicine (Approval Number: 23KT11). Written informed consent was obtained from the patient.

### Endomyocardial biopsy

An endomyocardial biopsy was performed to determine the cause of the patient’s sudden cardiac death. Using a disposable Cordis bioptome guided by fluoroscopy through the right femoral vein, six tissue fragments were obtained from the right ventricular septum. For immunohistochemical, Masson’s trichrome, and HE staining, tissue samples were fixed in formalin and embedded in paraffin, 5 μm thick sections were examined under light microscopy.

### Transmission electron microscopy

Myocardial tissue was pre-fixed with 1% glutaraldehyde followed by 1% osmium tetroxide fixation. The tissue was dehydrated step by step using a gradient ethanol solution, followed by gradient infiltration with a mixture of acetone and Epon812 resin and resin-embedded polymerization. The resin block was cut into 70 nm ultrathin sections, double-stained with uranyl acetate and lead citrate, and the specimen was observed under transmission electron microscopy (Talos L120C G2, Thermo Fisher Scientific) operating at 120 KeV. Images were recorded with a Ceta 4K x 4K CMOS camera.

### Focused ion beam scanning electron microscopy (FIB-SEM)

The resin block was observed under a dual beam scanning electron microscope (Aquilos Cryo-FIB, Thermo Fisher Scientific). Each serial face was imaged using T1 (in-lens detector) backscatter mode, with a 2.0 keV acceleration voltage and a current of 0.1 nA. The pixel size of X-Y panels was 5nm, and image dimension varied for each dataset (about 10-30μm). The resin block was sequentially milled by a gallium ion beam, with a Z-axis height of 10nm for each milling, and number of images varied for each dataset (about 800-1800). Images were aligned, cropped, and binned to form an image stack with 10nm isotropic resolution in X-Y-Z dimension for further segmentation and analysis.

### Viruses and mice

The BALB/c mice were purchased from Jinan Pengyue Experimental Animal Breeding Co. LTD. The SARS-CoV-2 variants Omicron BQ.1 and BA.5.2 were isolated from COVID-19 patients in Guangdong, China. Experiments related to authentic SARS-CoV-2 were conducted in Guangzhou Customs District Technology Center BSL-3 Laboratory.

### Animal experiments

6-8 weeks old mice were challenged with 1 × 10^5^ FFU SARS-CoV-2 (BQ.1 and BA.5.2). Mice were challenged with phosphate-buffered saline as controls. To investigate the ultrastructure of cardiomyocytes, hearts were collected seven days after infection. Myocardial tissue from mice was prepared using the same protocol as endomyocardial biopsy tissue.

### Proteome analysis

Mouse heart tissue was ground individually in liquid nitrogen and lysed with SDT lysis buffer, and protein concentration was determined by BCA method. Protein samples were hydrolyzed by trypsin, and desalination was conducted on the C18 desalting column, and peptides were collected and lyophilized. The peptides were dissolved and fractionated using a C18 column using a Rigol L3000 HPLC system. The eluates were separated and analyzed with an EvoSep One nano UPLC coupled to a Bruker timsTOF Pro2 mass spectrometry with a nano-electrospray ion source. The mass spectrometer adopts DDA PaSEF mode for data acquisition, and the full scan MS survey spectra range was m/z from 100 to 1700. The raw files were processed using SpectroMine software, MS spectra lists were searched against their species-level UniProt FASTA databases. Peptide identification was performed with an initial precursor mass deviation of up to 20 ppm and a fragment mass deviation of 20 ppm. Gene Ontology (GO) and InterPro (IPR) functional analysis were conducted using the interproscan program against the non-redundant protein database, and the databases of COG and KEGG were used to analyze the protein family and pathway.

## Data Availability

N/A

## Acknowledgements

This work was supported by National Natural Science Foundation of China (82070329 to ZL, 32241028 to ZL and PW, 82170521 to WC, 82170456 to YX, 82025001 to JZ, 82172240 to YW, 61932018 and 32241027 to FZ, 62072441 to XW).

## Author Contributions

Conceptualization, Z.L., Y.X., and W.C.; Methodology, Z.L., W.C., and Y.W.; Investigation, W.C., S.G., Y.W., X.W., B.T., H.L., J.A., M.Z., C.C., P.L., Z.Z., Y.W., X.H., X.W., J.Z., X.P., F.Z., P.W., J.Z., Y.X., and Z.L.; Resources, Z.L., Y.X., W.C. P.W., J.Z., Y.W., X.W., F.Z., and X.P.; Writing – Original Draft, Z.L., W.C., S.G., and Y.X; Writing – Review & Editing, Z.L.; Supervision, Z.L. and Y.X; Funding Acquisition, Z.L., W.C., Y.X., P.W., J.Z., Y.W., X.W., and F.Z.

## Declaration of Interests

The authors declare that they have no competing interests.

## Notes

### Competing Interest Statement

The authors have declared no competing interest.

### Author Declarations

Clinical study was approved by the Human Ethics Committee of Shanghai Tenth People?s Hospital, Tongji University School of Medicine (Approval Number: 23KT11). Written informed consent was obtained from the patient.

## REFERENCES

1. Adams, R.A., Liu, Z., Hsieh, C., Marko, M., Lederer, W.J., Jafri, M.S., and Mannella, C. (2023). Structural Analysis of Mitochondria in Cardiomyocytes: Insights into Bioenergetics and Membrane Remodeling. Current Issues in Molecular Biology 45, 6097–6115. 10.3390/cimb45070385.

2. Arnér, E.S., and Holmgren, A. (2000). Physiological functions of thioredoxin and thioredoxin reductase. Eur J Biochem 267, 6102–6109.

3. Baker, K.P., Schaniel, A., Vestweber, D., and Schatz, G. (1990). A yeast mitochondrial outer membrane protein essential for protein import and cell viability. Nature 348, 605–609.

4. Ballering, A.V., van Zon, S.K.R., Olde Hartman, T.C., and Rosmalen, J.G.M. (2022). Persistence of somatic symptoms after COVID-19 in the Netherlands: an observational cohort study. Lancet 400, 452–461.

5. Beverly, K.N., Sawaya, M.R., Schmid, E., and Koehler, C.M. (2008). The Tim8-Tim13 complex has multiple substrate binding sites and binds cooperatively to Tim23. J Mol Biol 382, 1144–1156.

6. Bian, W.P., Pu, S.Y., Xie, S.L., Wang, C., Deng, S., Strauss, P.R., and Pei, D.S. (2021). Loss of mpv17 affected early embryonic development via mitochondria dysfunction in zebrafish. Cell Death Discov 7, 250.

7. Billing, O., Kao, G., and Naredi, P. (2011). Mitochondrial function is required for secretion of DAF-28/insulin in C. elegans. PLoS One 6, e14507.

8. Brunk, U.T., and Terman, A. (2002). The mitochondrial-lysosomal axis theory of aging: accumulation of damaged mitochondria as a result of imperfect autophagocytosis. Eur J Biochem 269, 1996–2002.

9. Chaillou, T., Zhang, X., and McCarthy, J.J. (2016). Expression of Muscle-Specific Ribosomal Protein L3-Like Impairs Myotube Growth. J Cell Physiol 231, 1894–1902.

10. Conrad, M., Jakupoglu, C., Moreno, S.G., Lippl, S., Banjac, A., Schneider, M., Beck, H., Hatzopoulos, A.K., Just, U., Sinowatz, F., et al. (2004). Essential role for mitochondrial thioredoxin reductase in hematopoiesis, heart development, and heart function. Mol Cell Biol 24, 9414–9423.

11. Cunningham, K.S., Veinot, J.P., and Butany, J. (2006). An approach to endomyocardial biopsy interpretation. J Clin Pathol 59, 121–129.

12. Dalla Rosa, I., Cámara, Y., Durigon, R., Moss, C.F., Vidoni, S., Akman, G., Hunt, L., Johnson, M.A., Grocott, S., rWang, L., et al. (2016). MPV17 Loss Causes Deoxynucleotide Insufficiency and Slow DNA Replication in Mitochondria. PLoS Genet 12, e1005779.

13. Dallabona, C., Marsano, R.M., Arzuffi, P., Ghezzi, D., Mancini, P., Zeviani, M., Ferrero, I., and Donnini, C. (2010). Sym1, the yeast ortholog of the MPV17 human disease protein, is a stress-induced bioenergetic and morphogenetic mitochondrial modulator. Hum Mol Genet 19, 1098–1107.

14. Ding, C., Wu, Z., Huang, L., Wang, Y., Xue, J., Chen, S., Deng, Z., Wang, L., Song, Z., and Chen, S. (2015). Mitofilin and CHCHD6 physically interact with Sam50 to sustain cristae structure. Sci Rep 5, 16064.

15. Dolezal, P., Likic, V., Tachezy, J., and Lithgow, T. (2006). Evolution of the molecular machines for protein import into mitochondria. Science 313, 314–318.

16. Eisenstein, M. (2023). Seven technologies to watch in 2023. Nature 613, 794–797.

17. El’darov, C.M., Vays, V.B., Vangeli, I.M., Kolosova, N.G., and Bakeeva, L.E. (2015). Morphometric Examination of Mitochondrial Ultrastructure in Aging Cardiomyocytes. Biochemistry (Mosc) 80, 604–609.

18. Endo, T., and Kohda, D. (2002). Functions of outer membrane receptors in mitochondrial protein import. Biochim Biophys Acta 1592, 3–14.

19. Gallerne, C., Touat, Z., Chen, Z.X., Martel, C., Mayola, E., Sharaf el dein, O., Buron, N., Le Bras, M., Jacotot, E., Borgne-Sanchez, A., et al. (2010). The fourth isoform of the adenine nucleotide translocator inhibits mitochondrial apoptosis in cancer cells. Int J Biochem Cell Biol 42, 623–629.

20. Gluckman, T.J., Bhave, N.M., Allen, L.A., Chung, E.H., Spatz, E.S., Ammirati, E., Baggish, A.L., Bozkurt, B., Cornwell, W.K., 3rd, Harmon, K.G., et al. (2022). 2022 ACC Expert Consensus Decision Pathway on Cardiovascular Sequelae of COVID-19 in Adults: Myocarditis and Other Myocardial Involvement, Post-Acute Sequelae of SARS-CoV-2 Infection, and Return to Play: A Report of the American College of Cardiology Solution Set Oversight Committee. J Am Coll Cardiol 79, 1717–1756.

21. Gordon, D.E., Hiatt, J., Bouhaddou, M., Rezelj, V.V., Ulferts, S., Braberg, H., Jureka, A.S., Obernier, K., Guo, J.Z., Batra, J., et al. (2020). Comparative host-coronavirus protein interaction networks reveal pan-viral disease mechanisms. Science 370, eabe9403.

22. Guarnieri, J.W., Dybas, J.M., Fazelinia, H., Kim, M.S., Frere, J., Zhang, Y., Soto Albrecht, Y., Murdock, D.G., Angelin, A., Singh, L.N., et al. (2023). Core mitochondrial genes are down-regulated during SARS-CoV-2 infection of rodent and human hosts. Sci Transl Med 15, eabq1533.

23. Guo, T., Fan, Y., Chen, M., Wu, X., Zhang, L., He, T., Wang, H., Wan, J., Wang, X., and Lu, Z. (2020). Cardiovascular Implications of Fatal Outcomes of Patients With Coronavirus Disease 2019 (COVID-19). JAMA Cardiol 5, 811–818.

24. Harkness, T.A., Nargang, F.E., van der Klei, I., Neupert, W., and Lill, R. (1994). A crucial role of the mitochondrial protein import receptor MOM19 for the biogenesis of mitochondria. J Cell Biol 124, 637–648.

25. Helluy, X., Sauter, M., Ye, Y.X., Lykowsky, G., Kreutner, J., Yilmaz, A., Jahns, R., Boivin, V., Kandolf, R., Jakob, P.M., et al. (2017). In vivo T2* weighted MRI visualizes cardiac lesions in murine models of acute and chronic viral myocarditis. PLoS One 12, e0172084.

26. Huang, C., Wang, Y., Li, X., Ren, L., Zhao, J., Hu, Y., Zhang, L., Fan, G., Xu, J., Gu, X., et al. (2020). Clinical features of patients infected with 2019 novel coronavirus in Wuhan, China. Lancet 395, 497–506.

27. Humphries, A.D., Streimann, I.C., Stojanovski, D., Johnston, A.J., Yano, M., Hoogenraad, N.J., and Ryan, M.T. (2005). Dissection of the mitochondrial import and assembly pathway for human Tom40. J Biol Chem 280, 11535–11543.

28. Jiang, D., Zhao, L., and Clapham, D.E. (2009). Genome-wide RNAi screen identifies Letm1 as a mitochondrial Ca2+/H+ antiporter. Science 326, 144–147.

29. Kajihara, H., Taguchi, K., Hara, H., and Iijima, S. (1973). Electron microscopic observation of human hypertrophied myocardium. Acta Pathol Jpn 23, 335–347.

30. Kakimoto, Y., Okada, C., Kawabe, N., Sasaki, A., Tsukamoto, H., Nagao, R., and Osawa, M. (2019). Myocardial lipofuscin accumulation in ageing and sudden cardiac death. Sci Rep 9, 3304.

31. Kizilyaprak, C., Stierhof, Y.D., and Humbel, B.M. (2019). Volume microscopy in biology: FIB-SEM tomography. Tissue Cell 57, 123–128.

32. Koehler, C.M. (2004). The small Tim proteins and the twin Cx3C motif. Trends Biochem Sci 29, 1–4.

33. Kuroda, A., Hegab, A.E., Jingtao, G., Yamashita, S., Hizawa, N., Sakamoto, T., Yamada, H., Suzuki, S., Ishii, M., Namkoong, H., et al. (2017). Effects of the common polymorphism in the human aldehyde dehydrogenase 2 (ALDH2) gene on the lung. Respir Res 18, 69.

34. Lee, N.C., Dimmock, D., Hwu, W.L., Tang, L.Y., Huang, W.C., Chinault, A.C., and Wong, L.J. (2009). Simultaneous detection of mitochondrial DNA depletion and single-exon deletion in the deoxyguanosine gene using array-based comparative genomic hybridisation. Arch Dis Child 94, 55–58.

35. Liao, X., Ruan, X., Wu, X., Deng, Z., Qin, S., and Jiang, H. (2023). Identification of Timm13 protein translocase of the mitochondrial inner membrane as a potential mediator of liver fibrosis based on bioinformatics and experimental verification. J Transl Med 21, 188.

36. Liu, X., Ren, Z., Zhan, R., Wang, X., Wang, X., Zhang, Z., Leng, X., Yang, Z., and Qian, L. (2009). Prohibitin protects against oxidative stress-induced cell injury in cultured neonatal cardiomyocyte. Cell Stress Chaperones 14, 311–319.

37. Lu, G., Ling, Y., Jiang, M., Tan, Y., Wei, D., Jiang, L., Yu, S., Jiang, F., Wang, S., Dai, Y., et al. (2023). Primary assessment of the diversity of Omicron sublineages and the epidemiologic features of autumn/winter 2022 COVID-19 wave in Chinese mainland. Front Med 17, 758–767.

38. Lustbader, J.W., Cirilli, M., Lin, C., Xu, H.W., Takuma, K., Wang, N., Caspersen, C., Chen, X., Pollak, S., Chaney, M., et al. (2004). ABAD directly links Abeta to mitochondrial toxicity in Alzheimer’s disease. Science 304, 448–452.

39. Malkoff, D.B., and Strehler, B.L. (1963). The ultrastructure of isolated and in situ human cardiac age pigment. J Cell Biol 16, 611–616.

40. Mall, G., Mattfeldt, T., and Volk, B. (1980). Ultrastructural morphometric study on the rat heart after chronic ethanol feeding. Virchows Arch A Pathol Anat Histol 389, 59–77.

41. Mandel, H., Hartman, C., Berkowitz, D., Elpeleg, O.N., Manov, I., and Iancu, T.C. (2001). The hepatic mitochondrial DNA depletion syndrome: ultrastructural changes in liver biopsies. Hepatology 34, 776–784.

42. Maron, B.J., Udelson, J.E., Bonow, R.O., Nishimura, R.A., Ackerman, M.J., Estes, N.A.M., 3rd, Cooper, L.T., Jr., Link, M.S., and Maron, M.S. (2015). Eligibility and Disqualification Recommendations for Competitive Athletes With Cardiovascular Abnormalities: Task Force 3: Hypertrophic Cardiomyopathy, Arrhythmogenic Right Ventricular Cardiomyopathy and Other Cardiomyopathies, and Myocarditis: A Scientific Statement From the American Heart Association and American College of Cardiology. J Am Coll Cardiol 66, 2362–2371.

43. Merkwirth, C., Dargazanli, S., Tatsuta, T., Geimer, S., Löwer, B., Wunderlich, F.T., von Kleist-Retzow, J.C., Waisman, A., Westermann, B., and Langer, T. (2008). Prohibitins control cell proliferation and apoptosis by regulating OPA1-dependent cristae morphogenesis in mitochondria. Genes Dev 22, 476–488.

44. Nalbandian, A., Sehgal, K., Gupta, A., Madhavan, M.V., McGroder, C., Stevens, J.S., Cook, J.R., Nordvig, A.S., Shalev, D., Sehrawat, T.S., et al. (2021). Post-acute COVID-19 syndrome. Nat Med 27, 601–615.

45. Ott, C., Dorsch, E., Fraunholz, M., Straub, S., and Kozjak-Pavlovic, V. (2015). Detailed analysis of the human mitochondrial contact site complex indicate a hierarchy of subunits. PLoS One 10, e0120213.

46. Ott, C., Ross, K., Straub, S., Thiede, B., Götz, M., Goosmann, C., Krischke, M., Mueller, M.J., Krohne, G., Rudel, T., and Kozjak-Pavlovic, V. (2012). Sam50 functions in mitochondrial intermembrane space bridging and biogenesis of respiratory complexes. Mol Cell Biol 32, 1173–1188.

47. Peddie, C.J., Genoud, C., Kreshuk, A., Meechan, K., Micheva, K.D., Narayan, K., Pape, C., Parton, R.G., Schieber, N.L., Schwab, Y., et al. (2022). Volume electron microscopy. Nature Reviews Methods Primers 2, 51.

48. Petrakis, T.G., Ktistaki, E., Wang, L., Eriksson, S., and Talianidis, I. (1999). Cloning and characterization of mouse deoxyguanosine kinase. Evidence for a cytoplasmic isoform. J Biol Chem 274, 24726–24730.

49. Phelan, D., Kim, J.H., and Chung, E.H. (2020). A Game Plan for the Resumption of Sport and Exercise After Coronavirus Disease 2019 (COVID-19) Infection. JAMA Cardiol 5, 1085–1086.

50. Qiu, J., Wenz, L.S., Zerbes, R.M., Oeljeklaus, S., Bohnert, M., Stroud, D.A., Wirth, C., Ellenrieder, L., Thornton, N., Kutik, S., et al. (2013). Coupling of mitochondrial import and export translocases by receptor-mediated supercomplex formation. Cell 154, 596–608.

51. Rauschenberger, K., Schöler, K., Sass, J.O., Sauer, S., Djuric, Z., Rumig, C., Wolf, N.I., Okun, J.G., Kölker, S., Schwarz, H., et al. (2010). A non-enzymatic function of 17beta-hydroxysteroid dehydrogenase type 10 is required for mitochondrial integrity and cell survival. EMBO Mol Med 2, 51–62.

52. Richter-Dennerlein, R., Korwitz, A., Haag, M., Tatsuta, T., Dargazanli, S., Baker, M., Decker, T., Lamkemeyer, T., Rugarli, E.I., and Langer, T. (2014). DNAJC19, a mitochondrial cochaperone associated with cardiomyopathy, forms a complex with prohibitins to regulate cardiolipin remodeling. Cell Metab 20, 158–171.

53. Sagar, S., Liu, P.P., and Cooper, L.T., Jr. (2012). Myocarditis. Lancet 379, 738–747.

54. Seferović, P.M., Tsutsui, H., McNamara, D.M., Ristić, A.D., Basso, C., Bozkurt, B., Cooper, L.T., Jr., Filippatos, G., Ide, T., Inomata, T., et al. (2021). Heart Failure Association of the ESC, Heart Failure Society of America and Japanese Heart Failure Society Position statement on endomyocardial biopsy. Eur J Heart Fail 23, 854–871.

55. Shang, J., Ye, G., Shi, K., Wan, Y., Luo, C., Aihara, H., Geng, Q., Auerbach, A., and Li, F. (2020). Structural basis of receptor recognition by SARS-CoV-2. Nature 581, 221–224.

56. Signorile, A., Sgaramella, G., Bellomo, F., and De Rasmo, D. (2019). Prohibitins: A Critical Role in Mitochondrial Functions and Implication in Diseases. Cells 8.

57. Skrzypiec-Spring, M., Haczkiewicz, K., Sapa, A., Piasecki, T., Kwiatkowska, J., Ceremuga, I., Wozniak, M., Biczysko, W., Kobierzycki, C., Dziegiel, P., et al. (2018). Carvedilol Inhibits Matrix Metalloproteinase-2 Activation in Experimental Autoimmune Myocarditis: Possibilities of Cardioprotective Application. J Cardiovasc Pharmacol Ther 23, 89–97.

58. Skrzypiec-Spring, M., Sapa-Wojciechowska, A., Haczkiewicz-Leśniak, K., Piasecki, T., Kwiatkowska, J., Podhorska-Okołów, M., and Szeląg, A. (2021). HMG-CoA Reductase Inhibitor, Simvastatin Is Effective in Decreasing Degree of Myocarditis by Inhibiting Metalloproteinases Activation. Biomolecules 11, 1415.

59. Taanman, J.W., Bodnar, A.G., Cooper, J.M., Morris, A.A., Clayton, P.T., Leonard, J.V., and Schapira, A.H. (1997). Molecular mechanisms in mitochondrial DNA depletion syndrome. Hum Mol Genet 6, 935–942.

60. Tamai, S., Iida, H., Yokota, S., Sayano, T., Kiguchiya, S., Ishihara, N., Hayashi, J., Mihara, K., and Oka, T. (2008). Characterization of the mitochondrial protein LETM1, which maintains the mitochondrial tubular shapes and interacts with the AAA-ATPase BCS1L. J Cell Sci 121, 2588–2600.

61. Vercellino, I., and Sazanov, L.A. (2022). The assembly, regulation and function of the mitochondrial respiratory chain. Nat Rev Mol Cell Biol 23, 141–161.

62. Viscomi, C., Spinazzola, A., Maggioni, M., Fernandez-Vizarra, E., Massa, V., Pagano, C., Vettor, R., Mora, M., and Zeviani, M. (2009). Early-onset liver mtDNA depletion and late-onset proteinuric nephropathy in Mpv17 knockout mice. Hum Mol Genet 18, 12–26.

63. Wang, D., Hu, B., Hu, C., Zhu, F., Liu, X., Zhang, J., Wang, B., Xiang, H., Cheng, Z., Xiong, Y., et al. (2020). Clinical Characteristics of 138 Hospitalized Patients With 2019 Novel Coronavirus-Infected Pneumonia in Wuhan, China. Jama 323, 1061–1069.

64. Wang, X., Cai, B., Jia, Z., Chen, Y., Guo, S., Liu, Z., Wan, X., Zhang, F., and Hu, B. (2024). MitoStructSeg: A Comprehensive Platform for Mitochondrial Structure Segmentation and Analysis. bioRxiv 2024, 601295.

65. Wu, N.N., and Ren, J. (2019). Aldehyde Dehydrogenase 2 (ALDH2) and Aging: Is There a Sensible Link? Adv Exp Med Biol 1193, 237–253.

66. Xie, Y., Xu, E., Bowe, B., and Al-Aly, Z. (2022). Long-term cardiovascular outcomes of COVID-19. Nat Med 28, 583–590.

67. Xu, R., Kang, L., Wei, S., Yang, C., Fu, Y., Ding, Z., and Zou, Y. (2021). Samm50 Promotes Hypertrophy by Regulating Pink1-Dependent Mitophagy Signaling in Neonatal Cardiomyocytes. Front Cardiovasc Med 8, 748156.

68. Yang, J., Chen, T., and Zhou, Y. (2021). Mediators of SARS-CoV-2 entry are preferentially enriched in cardiomyocytes. Hereditas 158, 4.

69. Zeitlow, K., Charlambous, L., Ng, I., Gagrani, S., Mihovilovic, M., Luo, S., Rock, D.L., Saunders, A., Roses, A.D., and Gottschalk, W.K. (2017). The biological foundation of the genetic association of TOMM40 with late-onset Alzheimer’s disease. Biochim Biophys Acta Mol Basis Dis 1863, 2973–2986.

70. Zhang, X.M., Li, Y.C., Chen, P., Ye, S., Xie, S.H., Xia, W.J., and Yang, J.H. (2020). MG-132 attenuates cardiac deterioration of viral myocarditis via AMPK pathway. Biomed Pharmacother 126, 110091.

71. Zwacka, R.M., Reuter, A., Pfaff, E., Moll, J., Gorgas, K., Karasawa, M., and Weiher, H. (1994). The glomerulosclerosis gene Mpv17 encodes a peroxisomal protein producing reactive oxygen species. Embo j 13, 5129–5134.

