## Supplementary Information for "SARS-CoV-2 Damages Cardiomyocytes Mitochondria and Implicates Long COVID-associated Cardiovascular Manifestations"

[7] Lead contact

**Supplemental Table 1. Laboratory Results for Patient #1 (out of range data are shown in ed). Related to Figure 1.**

| | Value | Normal Value or Range |
|---|---|---|
| **Inflammatory markers** | | |
| C-reactive protein (mg/L) | 1.18 | < 8.2 |
| Plasma D-dimer (mg/L) | 1.98 | < 0.55 |
| **Cardiac biomarkers** | | |
| Troponin T (ng/ml) | 0.30 | < 0.014 |
| Creatine kinase-MB (ng/ml) | 7.21 | 0.1-2.88 |
| Myoglobin (μg/L) | 101.90 | 20-80 |
| N-Terminal pro-brain natriuretic peptide (pg/ml) | 54.01 | < 125 |
| **Hepatic function panel** | | |
| Albumin (g/L) | 42.10 | 40-55 |
| Serum conjugated bilirubin (μmol/L) | 1.00 | 0-5.0 |
| Serum total bilirubin (μmol/L) | 9.50 | 3.4-17.1 |
| Alanine transaminase (U/L) | 141.00 | 7-40 |
| Aspartate transaminase (U/L) | 115.30 | 13-35 |
| **Basic metabolic panel** | | |
| Serum creatinine (μmol/L) | 80.90 | 45-84 |
| Blood uric acid (μmol/L) | 415.50 | 135-357 |
| Blood glucose (μmol/L) | 6.69 | 3.9-6.1 |
| Total cholesterol (mmol/L） | 4.11 | 3.6-5.2 |
| Triglycerides (mmol/L) | 0.75 | 0.57-1.71 |
| Potassium (mmol/L) | 3.65 | 3.5-5.3 |
| Sodium (mmol/L) | 140.00 | 137-147 |
| Chloride (mmol/L) | 103.00 | 99-110 |
| **Complete blood count** | | |
| White blood cell count (×$10^9$/L) | 9.18 | 3.5-9.5 |
| Hemoglobin (g/L) | 145.00 | 115-150 |
| Hematocrit (%) | 43.20 | 35-45 |
| Platelets (×$10^9$/L) | 157.00 | 125-350 |
| Percentage of neutrophils (%) | 83.20 | 40-75 |
| Percentage of lymphocyte (%) | 10.60 | 20-50 |
| Neutrophil differential count (×$10^9$/L) | 7.63 | 1.8-6.3 |
| **Coagulation profile** | | |
| Prothrombin time (s) | 11.00 | 9.0-15.0 |
| International normalization ratio | 0.92 | 1.5-2.8 |
| Activated partial thromboplastin time (s) | 19.60 | 19.9-39.9 |
| **Molecular biology profile** | | |
| SARS-CoV-2 nucleic acid | Negative | Negative |
| **Immunological panel** | | |
| Coxsackie group B virus IgM | Negative | Negative |
| Adenovirus IgM | Negative | Negative |
| Respiratory syncytial virus IgM | Negative | Negative |
| Rheumatoid factor (IU/mL) | <11.6 | <15.8 |
| P-ANCA (U/mL) | Negative | Negative |

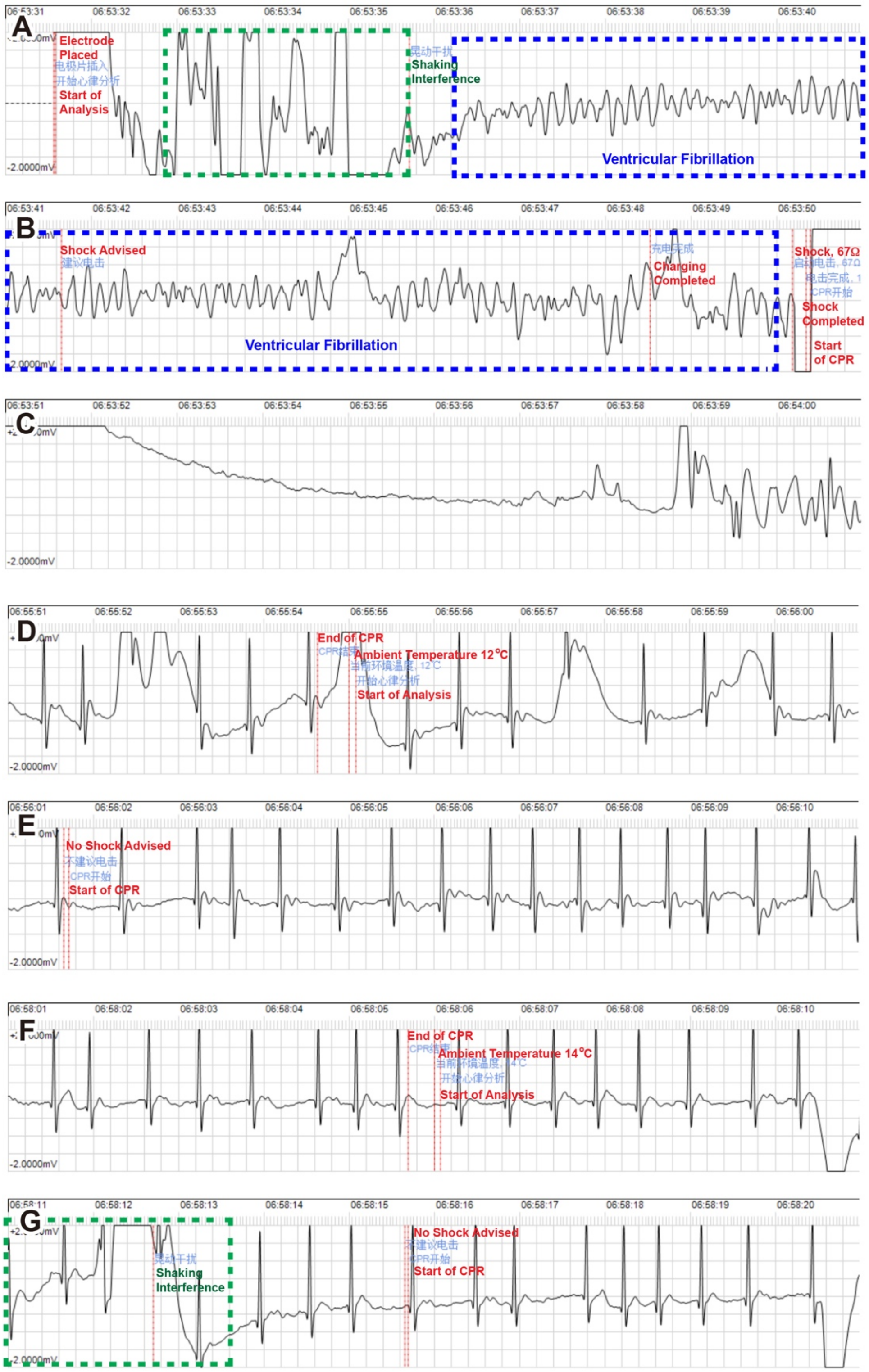


**Figure S1. ECG recordings retrieved from the AED. Related to Figure 1.**

Blue frames in panel A and B indicated coarse ventricular fibrillation before the shock. Green frames in panel A and G are shaking interference.

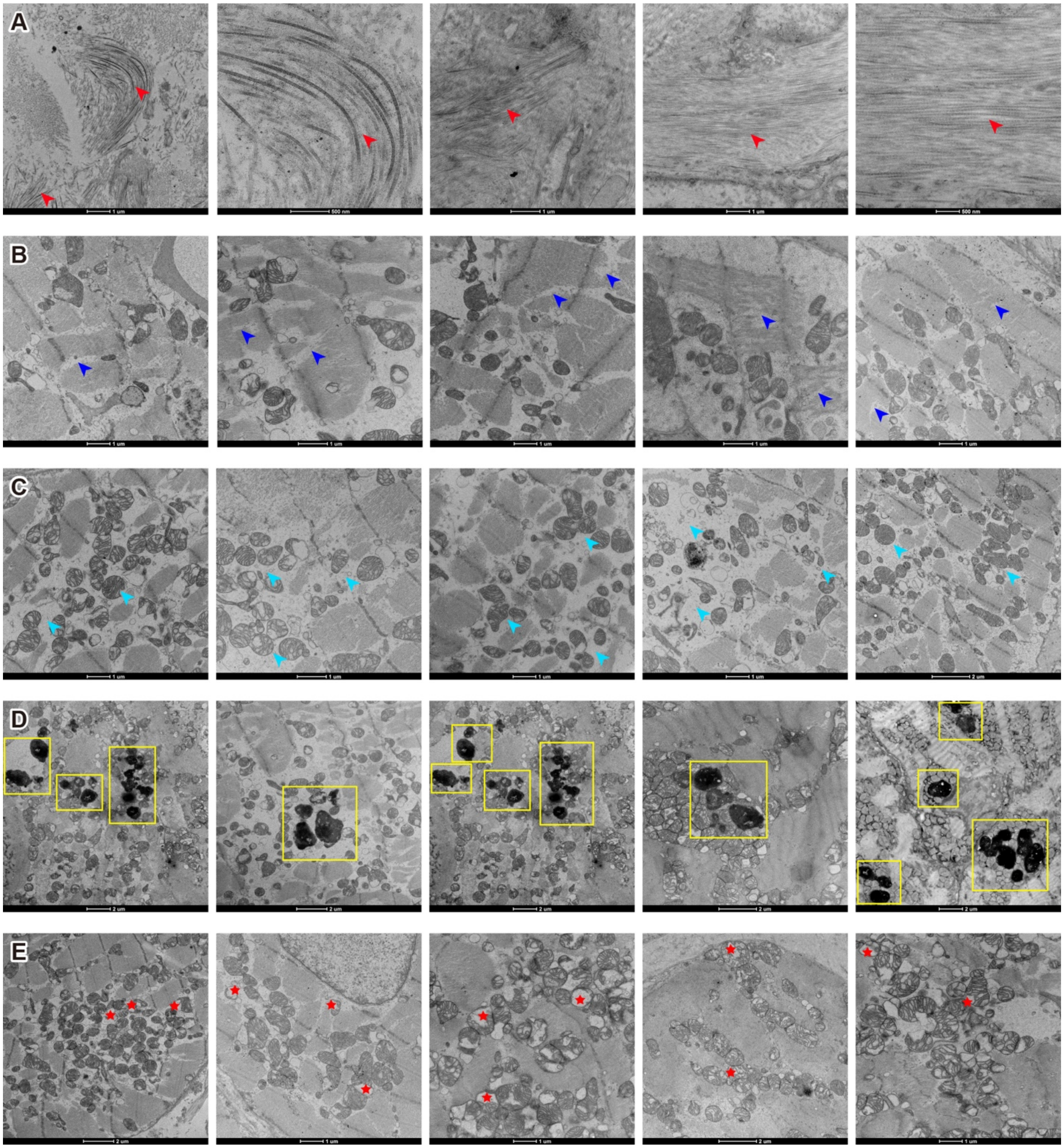


**Figure S2. Electron micrographs of endomyocardial biopsy tissue sample from patient #1. Related to Figure 1.**

Row A, red arrowheads indicated interstitial collagen fiber deposits. Row B, blue arrowheads indicated loss of integrity of myofibrillar bundles, including broken and disordered myofibrils, Z-lines broken, loss of sarcomere, and filling of mitochondria. Row

C, cyan arrowheads indicated necrosis of myofibrillar bundles, with severe loss of sarcomere and replacement with mitochondria. Row D, yellow frames indicated lipofuscin granules accumulation. Row E, red stars indicated swollen and vacuolated mitochondria, with distorted and broken cristae.

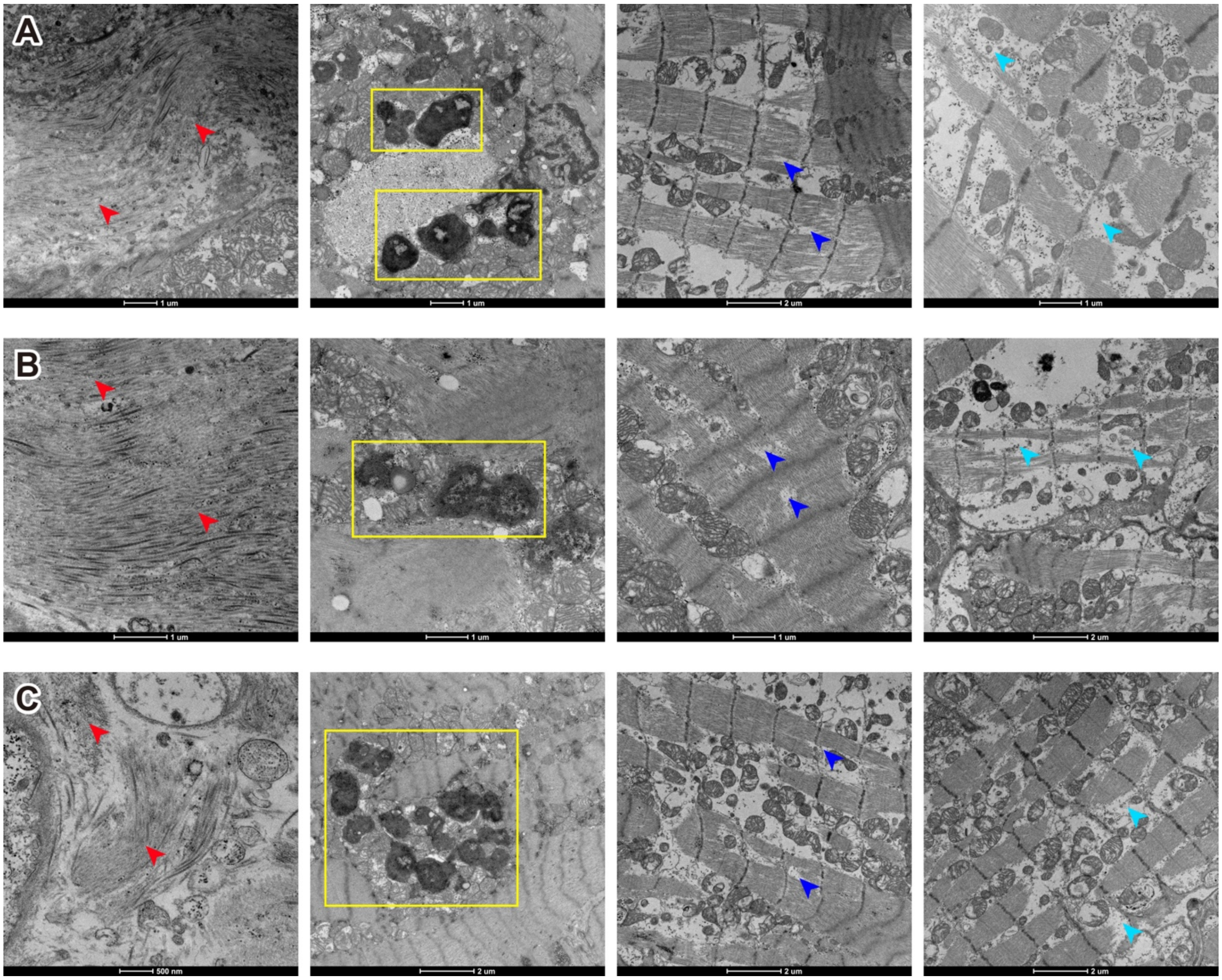

**Figure S3. Electron micrographs of endomyocardial biopsy tissue sample from patient #3, #4, and #5. Related to Figure 3.**

Row A-C, electron micrographs of endomyocardial biopsy tissue sample from patient #3-#5, respectively. Red arrowheads indicated interstitial collagen fiber deposits; yellow frames indicated lipofuscin granules accumulation; blue arrowheads indicated loss of integrity of myofibrillar bundles; cyan arrowheads indicated necrosis of myofibrillar bundles.
